# Design and preliminary safety validation of a hybrid deterministic–AI triage system for multilingual primary healthcare: a WhatsApp-based vignette study in South Africa

**DOI:** 10.64898/2026.04.21.26349781

**Authors:** Bongekile Esther Nkosi-Mjadu

## Abstract

**Background:** South Africa’s public healthcare system serves most of the population through approximately 3,900 primary healthcare clinics characterised by long waiting times and high volumes of repeat-prescription visits. No published pre-arrival digital triage system operates across all 11 official South African languages while aligning with the South African Triage Scale (SATS). This paper reports the design and preliminary safety validation of BIZUSIZO, a hybrid deterministic-AI WhatsApp triage system.

**Methods:** BIZUSIZO delivers SATS-aligned triage via WhatsApp, combining AI-assisted free-text classification (Claude Haiku 4.5) with a Deterministic Clinical Safety Layer (DCSL) that overrides AI output for 53 clinical discriminator categories (14 RED, 19 ORANGE, 20 YELLOW) coded in all 11 official languages and independent of AI availability. A five-domain risk factor assessment can only upgrade triage level. One hundred and twenty clinical vignettes in patient language (English, isiZulu, isiXhosa, Afrikaans; 30 per language) were scored against a developer-assigned gold standard with independent blinded nurse review. A 121-vignette multilingual DCSL safety consistency check across all 11 languages and a 220-call post-hoc framing sensitivity evaluation (110 paired vignettes) were also conducted.

**Results:** Under-triage was 3.3% (4/120; 95% CI: 0.9%–8.3%) with no RED under-triage; exact concordance was 80.0% (96/120) and quadratic weighted kappa 0.891 (95% CI: 0.827–0.932). One two-level under-triage was observed on a non-RED presentation (V072, isiXhosa burns vignette, ORANGE⍰GREEN); one two-level over-triage was observed (V054, isiZulu deep laceration, YELLOW⍰RED). In the framing sensitivity evaluation, AI-only classification achieved 50.9% RED invariance under adversarial framing; full-pipeline classification achieved 95.0% in four validated languages, with the DCSL rescuing 18 of 23 AI drift cases.

**Conclusions:** A hybrid deterministic-AI triage system with DCSL-based emergency detection achieved zero RED under-triage and consistent RED detection across all 11 official languages. The 16.7% over-triage rate falls within published South African SATS ranges (13.1–49%). A single two-level under-triage event was observed on an isiXhosa burns vignette (ORANGE⍰GREEN) and is discussed in Limitations. Findings are preliminary; prospective validation against independent nurse triage is the necessary next step.

## Background

South Africa’s public healthcare system faces a structural mismatch between patient demand and facility capacity. Approximately 83% of the South African population depends on the public healthcare sector [23], which delivers primary care through approximately 3,900 clinics. Waiting times are persistently long: a before-and-after evaluation of Ideal Clinic system-strengthening interventions across nine clinics in KwaZulu-Natal reported a median overall facility time of 122 minutes (IQR 81–204) post-intervention, with 32% of patients spending more than 3 hours during their visit and only 6% of total time spent in direct clinical consultation [24]. Clinics are described by frontline providers as engaged in ‘pushing numbers’ rather than individualised clinical care [1]. Up to 70% of a facility’s daily prescription load involves the preparation of repeat prescriptions for stable chronic patients [25], and in the same KwaZulu-Natal evaluation a combined 50% of all patients seen at baseline were attending for chronic non-communicable disease, antiretroviral therapy, or tuberculosis services [24]. The national Central Chronic Medicine Dispensing and Distribution (CCMDD) programme, established to reduce this burden, now actively services over 2 million patients across 3,436 facilities in 46 districts [3], and the Ideal Clinic framework requires pre-dispensed medication or CCMDD enrolment for clinically stable chronic patients [2]. Despite these mechanisms, the healthcare system still lacks a way to evaluate and direct patient demand before physical arrival at a facility – a routing failure at the point of entry.

The CCMDD programme, South Africa’s primary differentiated care model for chronic medication distribution, had scaled to over 2 million actively serviced patients across 3,436 health facilities in 46 health districts by October 2019 and has continued to expand since [3]. Despite its scale, patient adherence remains a persistent challenge: a pill-count study of HIV-positive CCMDD enrollees in the Eastern Cape found that 56.6% had sub-optimal adherence (below the 95% threshold), with reasons including medication unavailability, adverse effects, and regimen complexity [4]. These data suggest that the absence of a digital navigation and reminder layer between patients and the care system contributes to facility overcrowding and suboptimal patient flow.

The National Health Insurance (NHI) Act 20 of 2023 establishes the policy framework for universal health coverage and contemplates an integrated, coordinated national health system (Section 32) supported by a national health information platform (Section 34) [5]. This legislative framework creates a policy environment aligned with digital health tools that can help route demand, reduce unnecessary facility visits, and improve emergency response for patients who genuinely require urgent care.

Several digital health tools have been deployed in African and global healthcare settings, including MomConnect, South Africa’s national maternal health messaging service, which had reached over 1.5 million pregnant women by 2018 and has continued to scale since [6], and the World Health Organization’s Health Alert chatbot deployed globally during the COVID-19 pandemic [7]. Internationally, AI-assisted triage systems have shown promise in emergency department settings [8]. However, no published system provides the combination of SATS-aligned clinical triage, all 11 South African official languages, deterministic safety overrides independent of artificial intelligence, clinical governance with defined safety targets, and integration with facility routing in a single WhatsApp-based platform.

The South African Triage Scale is a validated triage tool used in hospital emergency departments across the country. SATS employs a physiologically based Triage Early Warning Score (TEWS) together with clinical discriminators to classify patients into four urgency categories: RED (emergency), ORANGE (very urgent), YELLOW (urgent), and GREEN (routine) [9]. While SATS has been validated for in-hospital use by trained clinicians with access to vital signs and physical examination, its performance differs substantially outside the hospital setting. Mould-Millman et al. found that prehospital emergency medical service (EMS) providers using SATS achieved only 56.5% concordance with gold standard triage, with an under-triage rate of 29.5%, primarily attributable to difficulties with clinical discriminators and TEWS calculation in the field [10]. Digital triage via WhatsApp is analogous to prehospital triage: the patient reports symptoms remotely, without physical examination, vital sign measurement, or visual assessment. This limitation means a digital triage system must compensate for missing clinical data with conservative classification.

Recent evidence has further underscored the gap between controlled evaluation and real-world deployment of AI in clinical settings. Bean et al. conducted a randomised study with 1,298 participants and found that while large language models achieved 94.9% condition identification under simulated conditions, accuracy fell below 34.5% when real users interacted with the systems – worse than the control group using conventional search – primarily because users provided incomplete symptom information [22]. Yun et al. further demonstrated that LLMs exhibit systematic sensitivity to question framing across 6,614 clinical query pairs and eight language models, with positively and negatively framed queries about identical clinical evidence producing contradictory model conclusions and multi-turn conversations amplifying the effect [21]. These two findings – incomplete real-world input and framing-driven classification drift – are not peripheral failure modes but foundational design constraints for any AI-assisted triage system intended for deployment to self-presenting patients. Framing vulnerability in particular cannot be resolved through prompt engineering or model selection alone; it requires architectural containment. The safety architecture described below was designed with both failure modes as primary design inputs, not as post-hoc mitigations, and the system was evaluated against both through dedicated test suites (symptom completeness handling, runtime minimisation detection, and a 220-call framing sensitivity evaluation reported in Results).

This paper reports the design and preliminary safety validation of BIZUSIZO, a hybrid deterministic-AI triage system delivered via WhatsApp for South African primary healthcare, including its clinical methodology, safety architecture, and clinical governance framework. It reports the results of a 120-vignette validation study benchmarked against published SATS performance data, together with a post-hoc framing sensitivity evaluation. The paper does not report patient outcomes; a formal pilot concordance study is planned. This study makes three specific contributions to the digital health literature: (1) a hybrid deterministic-probabilistic safety architecture that combines AI-assisted triage with AI-independent emergency detection, designed for deployment resilience in resource-constrained settings; (2) a multilingual safety design framework extending SATS-aligned triage to all 11 official South African languages, with explicit treatment of code-switching, patient-language terminology, and culturally specific safety mechanisms including runtime minimisation detection; and (3) a continuous integration clinical safety pipeline that uses an automated regression test suite as a deployment gate, operationalising evaluation as safety infrastructure rather than quality assurance. Underlying all three contributions is a reframing of the core problem: AI safety in resource-constrained primary healthcare is most productively treated as a systems engineering problem – solved through layered deterministic constraints, multi-agent verification, and continuous monitoring – rather than a model accuracy problem. This framing has direct implications for how AI-assisted clinical tools should be designed, validated, and governed in low-resource settings where AI service reliability, language coverage, and clinical oversight capacity cannot be assumed.

## Methods

### System architecture and patient journey

BIZUSIZO operates on WhatsApp Business API. WhatsApp was selected because it requires no application download, functions on low-cost smartphones — a critical factor given that 96.1% of South African households own at least one mobile phone [11] — and is used by 68% of active instant messaging users in South Africa [12]. The backend comprises a Node.js server deployed on Railway (a cloud platform-as-a-service), a PostgreSQL database hosted on Supabase, and the Anthropic Claude family of large language models for AI-assisted free-text symptom classification. The 120-vignette validation reported in this paper was conducted using Claude Haiku 4.5 (claude-haiku-4-5-20251001) as the primary classifier. Following validation, language-specific ceiling effects identified during 121-vignette DCSL regression testing prompted an upgrade of the production classifier to Claude Sonnet 4 (claude-sonnet-4-20250514) prior to pilot deployment; the Sonnet 4 configuration has not been independently validated on a held-out vignette set and awaits formal validation as part of the planned prospective pilot (see Limitations).

The system architecture is presented in Figure 1. The patient interaction follows a structured sequence: (1) language selection from all 11 official South African languages (English, isiZulu, isiXhosa, Afrikaans, Sepedi, Setswana, Sesotho, Xitsonga, siSwati, Tshivenda, isiNdebele); (2) informed consent incorporating disclosure that AI may be used for symptom assessment, reference to the Protection of Personal Information Act, and the national emergency number (10177); (3) symptom input via either a structured 15-category menu, a voice note option, or free-text description; (4) triage classification through three parallel pathways; (5) five-domain risk factor assessment; (6) urgency-differentiated routing – RED patients are directed to emergency services, while ORANGE, YELLOW, and GREEN patients are routed via GPS to the nearest primary healthcare facility; (7) clinic consultation with nurse triage review and, where clinically indicated, formal hospital referral via a portable referral identifier; and (8) a two-stage automated follow-up comprising a 24-hour check-in and a 72-hour symptom assessment, with automatic triage upgrade for patients reporting worsening symptoms. Patients may withdraw consent at any time by sending ‘STOP,’ which terminates the session and logs the withdrawal.

**Figure 1.**
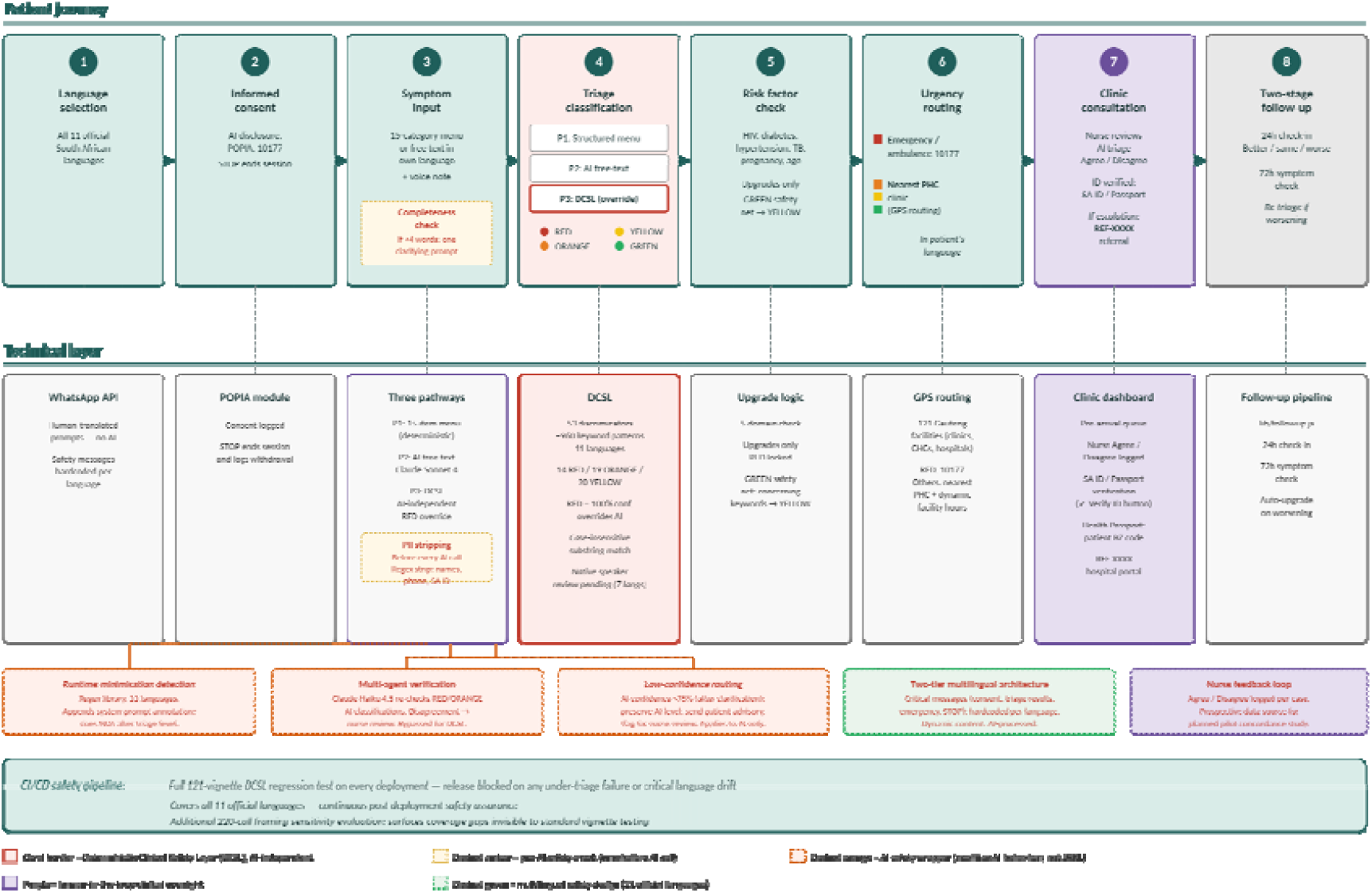
BIZUSIZO patient journey (top) and corresponding technical components (bottom). Step 3 (symptom input) is preceded by a symptom completeness check. PII stripping runs before every AI call and is shown as a technical-layer annotation within the Three pathways box. The Deterministic Clinical Safety Layer (DCSL) is shown with a coral border as one of three triage pathways at step 4 and operates independently of the AI classifier. Multi-agent verification is invoked for RED/ORANGE AI classifications; low-confidence AI classifications (<75%) are routed to nurse review. Step 6 shows urgency-differentiated routing: RED to emergency services, ORANGE/YELLOW/GREEN to GPS-routed primary healthcare facilities. Step 7 shows nurse review, identity verification, and the hospital referral pathway via portable REF-XXXX identifier. The CI/CD safety pipeline runs the full 121-vignette DCSL regression test across all 11 official languages on every deployment; release is blocked on any under-triage failure or critical language drift. A 220-call framing sensitivity evaluation surfaces coverage gaps invisible to standard vignette testing.

Triage routing is differentiated by urgency level. RED-classified patients receive an instruction to call an ambulance or proceed directly to the nearest emergency unit, with the national emergency number (10177) provided. ORANGE, YELLOW, and GREEN patients are routed via GPS to the nearest of 121 mapped public healthcare facilities in Gauteng, spanning clinics, community health centres, and hospitals. At pilot facilities, a web-based clinic dashboard provides reception staff and triage nurses with a real-time pre-arrival queue sorted by triage colour, enabling file preparation before the patient arrives. Six patient streams are aligned with national Department of Health primary healthcare standards: RED (emergency, immediate), ORANGE (fast-track, within 30 minutes), YELLOW and GREEN (routine), walk-in registration for patients who did not use WhatsApp triage, and chronic medication bypass for stable Central Chronic Medicine Dispensing and Distribution programme patients. Following consultation, the treating nurse records whether the AI-assigned triage level matched the clinical assessment (Agree) or required correction (Disagree); this structured feedback is logged with timestamp and staff identifier, and constitutes a prospective data source for the planned pilot concordance study. Where the nurse’s clinical assessment indicates the patient requires hospital-level care, a formal escalation pathway is available: the nurse records the clinical reason and selects transport method (ambulance or self-transport), and the system generates a portable referral identifier (REF-XXXX) that is sent to the patient’s WhatsApp and is retrievable by the receiving hospital via a dedicated lookup portal. This referral architecture is designed to operate within South Africa’s district health system model, in which primary healthcare facilities serve as the first point of care and hospital referral is a clinical decision made by the treating nurse rather than an automated system output.

### Clinical methodology: SATS alignment

Fifteen symptom categories are available via the structured menu: breathing/chest pain, head injury, pregnancy-related, bleeding, fever/cough, stomach/vomiting, child illness, medication/chronic, bone/joint pain, mental health, allergy/rash, women’s health (family planning), health screening (HIV, blood pressure, diabetes), other, or request to speak to a human. Thirteen of these were mapped to SATS clinical discriminators and organised into three triage pathways; the women’s health and health screening categories route to nurse-led service delivery rather than urgency triage. The first triage pathway uses the structured menu where patients select from the 13 clinically-mapped symptom categories, producing deterministic triage assignments without AI involvement.

The second pathway processes free-text symptom descriptions through Claude Haiku 4.5 (Anthropic), a large language model which classifies the input into RED, ORANGE, YELLOW, or GREEN with an associated confidence score between 0 and 100. The AI system prompt instructs the model to classify in any of South Africa’s 11 official languages, recognise code-switching (mixing of languages common in South African speech) and township medical terminology (such as ‘sugar’ for diabetes or ‘high blood’ for hypertension), and classify upward when uncertain. The model returns a structured reasoning trace of up to 50 words explaining which SATS step determined the classification, along with an estimated Triage Early Warning Score proxy, both of which are logged for audit review. The governance framework specifies a 75% confidence threshold below which classifications are flagged for human clinical review; this escalation pathway is logged for audit and constitutes a planned feature of the pilot governance protocol.

The third pathway is the DCSL, which operates independently of AI and overrides any AI classification when recognised life-threatening symptom combinations are detected in the patient’s text (described below).

A five-domain risk factor assessment evaluates age, chronic conditions (HIV, hypertension, diabetes, tuberculosis), immunocompromised status, pregnancy, and functional status. This assessment can only upgrade a triage level, never downgrade. A RED classification is never reduced regardless of risk factor profile. This one-directional adjustment ensures that the system’s conservative bias compounds rather than cancels.

### Symptom completeness validation

Prior to triage classification, the system performs a symptom completeness check on all incoming patient messages. Messages containing fewer than four words and lacking recognised clinical symptom keywords (assessed via regular expression matching across English and indigenous South African language terms) are flagged as insufficiently detailed for safe triage. In such cases, rather than proceeding with classification on incomplete information, the system sends a single targeted follow-up question in the patient’s detected language, prompting them to specify symptom location, onset, and trajectory. The follow-up prompt is available in all 11 official South African languages. If the patient responds with additional detail, the original message and clarification are concatenated and processed as a single triage input. This mechanism was implemented in response to evidence from Bean et al. [22], who demonstrated that incomplete user input was the primary driver of real-world large language model triage failure, with diagnostic accuracy declining from 94.9% under structured vignette conditions to below 34.5% when users provided unstructured, insufficient symptom descriptions. This design directly addresses the gap between controlled evaluation conditions and real-world clinical deployment, where patients may describe their symptoms in as few as one or two words.

### Runtime minimisation detection

The system implements runtime detection of minimising language patterns across all 11 official South African languages. This feature addresses a culturally specific phenomenon in South African primary healthcare settings, where patients frequently downplay symptoms to avoid being perceived as troublesome or wasting clinical resources. The detection layer comprises a library of regular expression patterns covering semantically equivalent minimisation phrases in English, isiZulu, isiXhosa, Afrikaans, Sepedi, Setswana, Sesotho, Xitsonga, siSwati, Tshivenda, and isiNdebele. Pattern categories include expressions conveying that symptoms are “probably nothing” or “not that bad,” apologies for seeking care, claims of coping ability, and self-accusations of overreacting. When minimising language is detected, a system prompt annotation is appended to the AI triage request, instructing the model to weight clinical discriminators and objective indicators more heavily than the patient’s self-assessment of severity. This mechanism does not alter the triage classification directly; rather, it informs the AI model’s reasoning process by contextualising the patient’s language within known cultural health-seeking behaviour patterns. The minimisation detection patterns were developed through consultation with the clinical team but remain pending formal review by native speakers of each language to ensure linguistic accuracy and cultural appropriateness. To the authors’ knowledge, no previously published clinical AI triage system has implemented runtime detection of patient minimisation language as a safety mechanism (based on searches of PubMed and Google Scholar conducted through April 2026 using combinations of the terms “patient minimisation,” “symptom minimisation,” “AI triage,” “clinical decision support,” and “large language model triage”), and formal sensitivity and specificity assessment of the detection patterns is planned as part of prospective validation.

### Deterministic safety architecture

The clinical rules engine scans the patient’s raw symptom text for keyword combinations associated with 53 clinical discriminator categories across three triage levels: 14 RED (life-threatening), 19 ORANGE (very urgent), and 20 YELLOW (urgent) (Table 1). This component is referred to hereafter as the Deterministic Clinical Safety Layer (DCSL). BIZUSIZO’s overall architecture is a hybrid deterministic-probabilistic safety system: the DCSL provides AI-independent emergency detection, while the AI classifier handles probabilistic urgency grading for non-life-threatening presentations. When a match is detected, the DCSL assigns RED with 100% confidence, overriding any AI output. Keywords are expressed in natural patient language rather than clinical terminology. This design decision was informed by initial stress testing, which revealed three critical bugs: the phrase ‘can’t breathe’ (how patients describe respiratory distress) did not match the clinical term ‘shortness of breath’; the isiXhosa word for breathing (‘phefumla’) did not match the isiZulu variant (‘phefumuli’); and ‘bitten by a snake’ did not match the exact string ‘snake bite.’ After correction and expansion of the keyword vocabulary, the DCSL was retested and achieved 100% sensitivity (121/121) on the validation test suite across all coded languages, with no false positives observed within that suite. Real-world sensitivity remains untested and requires prospective validation against live patient presentations. Following this initial validation in four languages (English, isiZulu, isiXhosa, and Afrikaans), keyword coverage was subsequently extended to all 11 official South African languages prior to pilot deployment. A dedicated validation suite of 121 test cases (116 positive, 5 negative) passes on all cases across all 11 languages, with exit code 1 on any failure blocking deployment. Keyword accuracy for the seven additional languages awaits independent review by native speakers, particularly for Sepedi, Setswana, Xitsonga, Tshivenda, and isiNdebele, where colloquial usage patterns are less well-documented; this review is planned prior to pilot commencement.

**Table 1.**
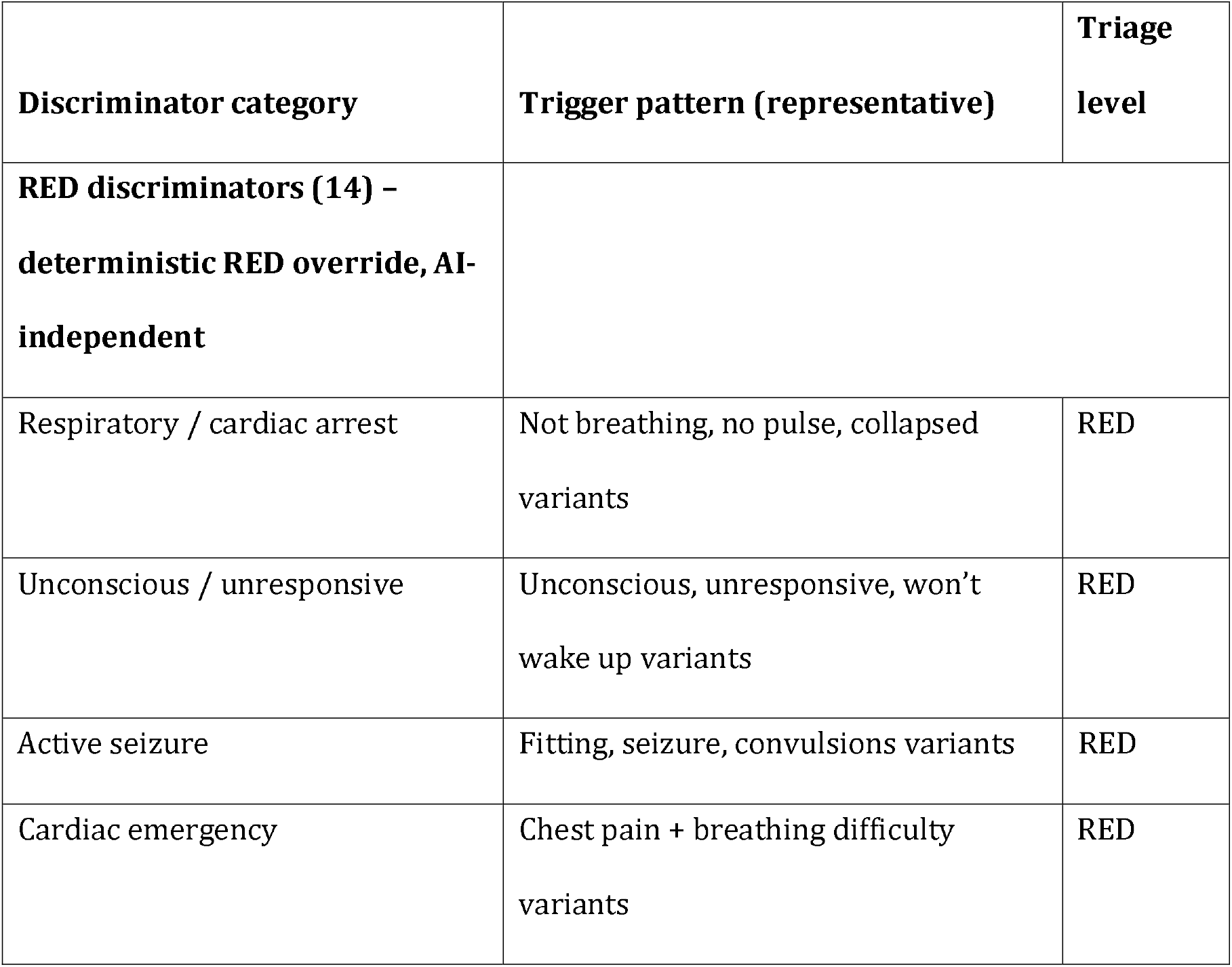
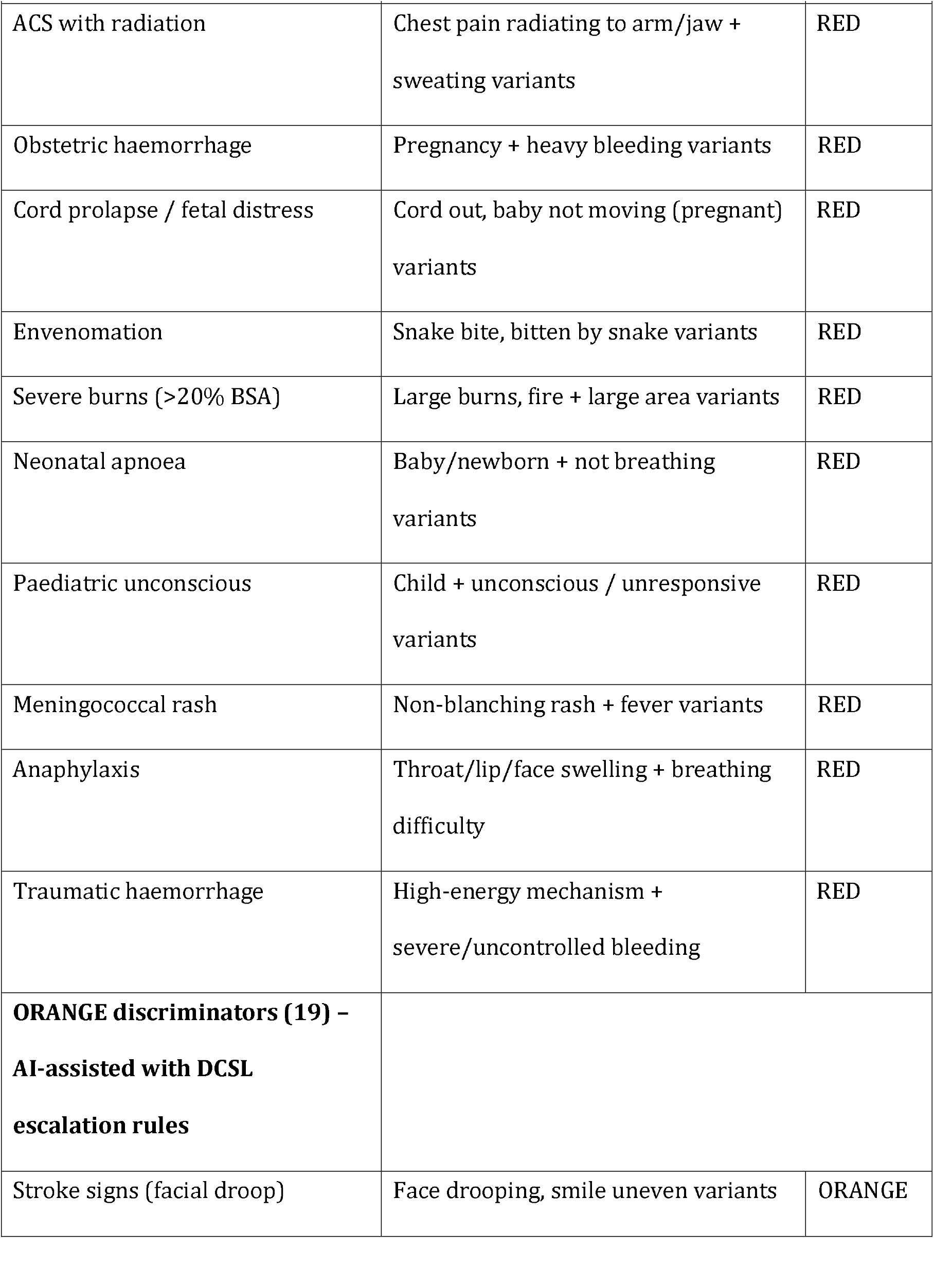
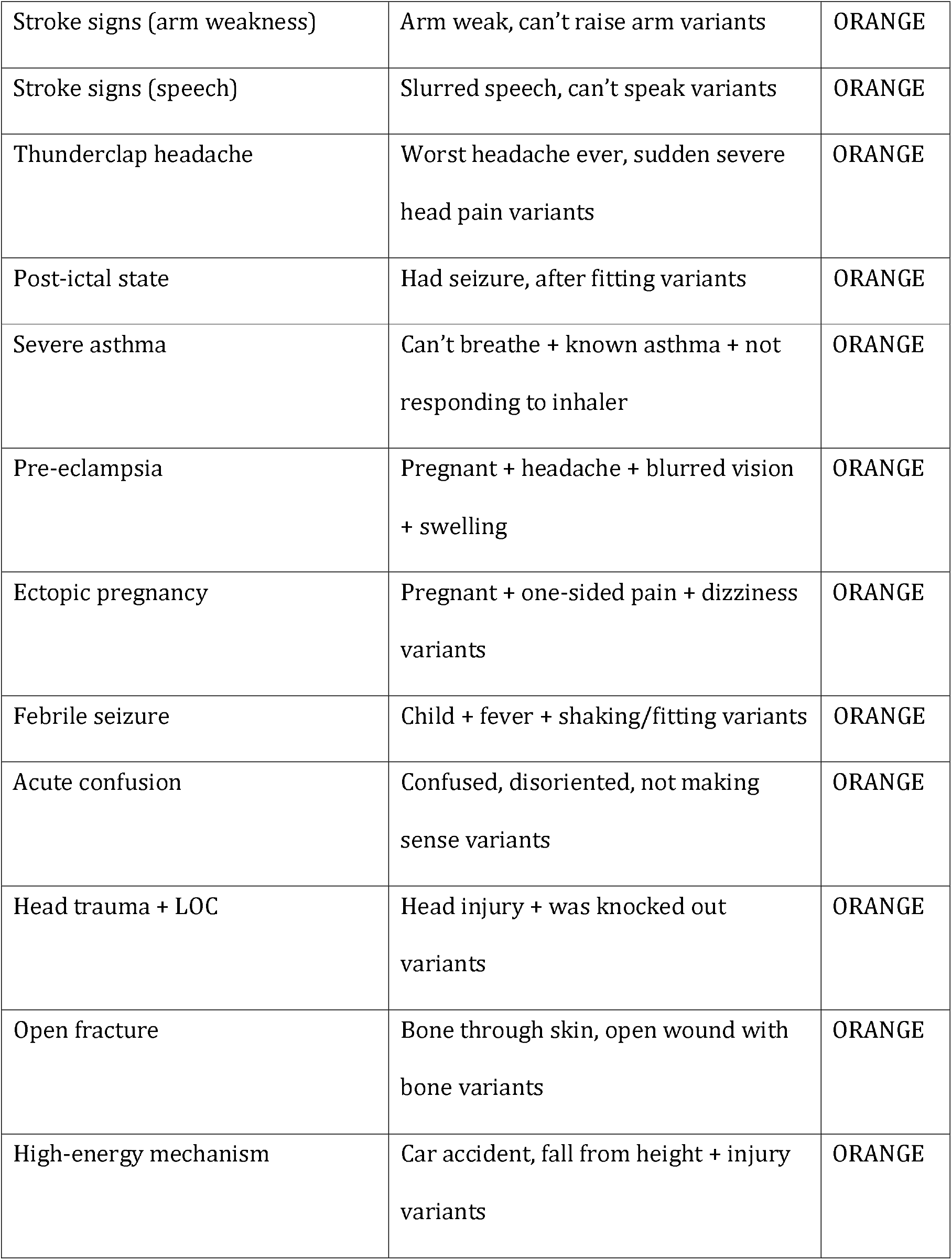
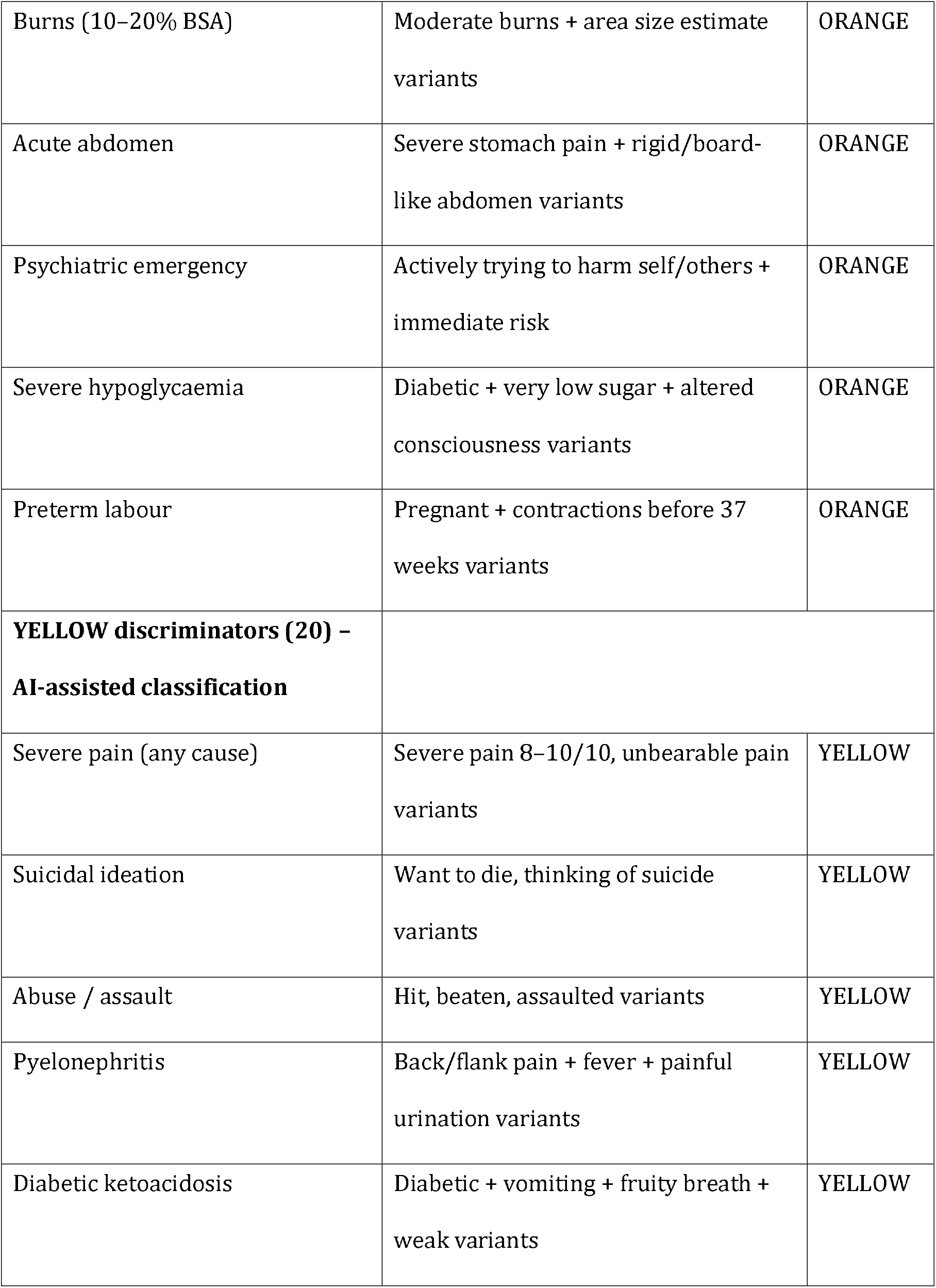
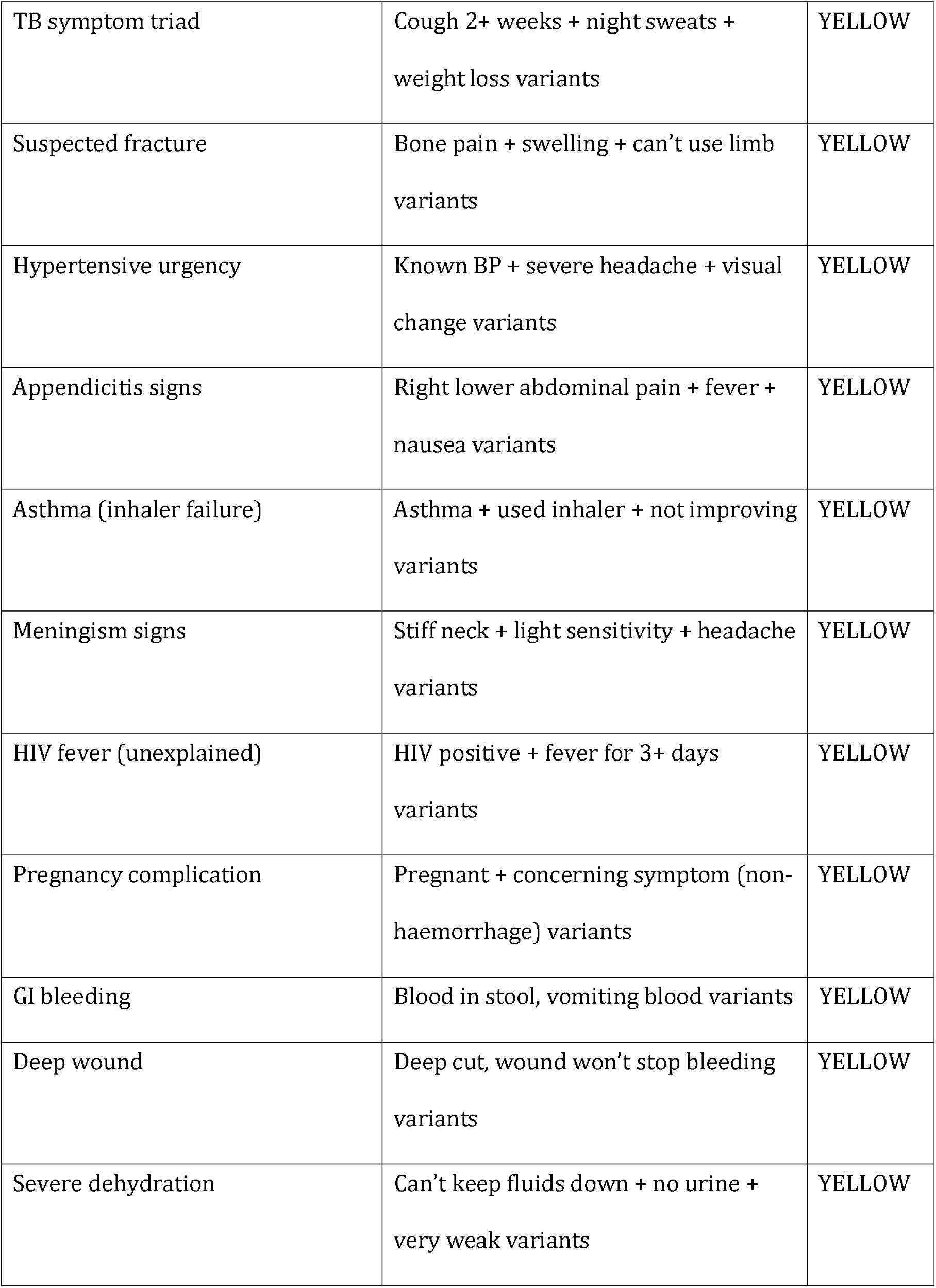
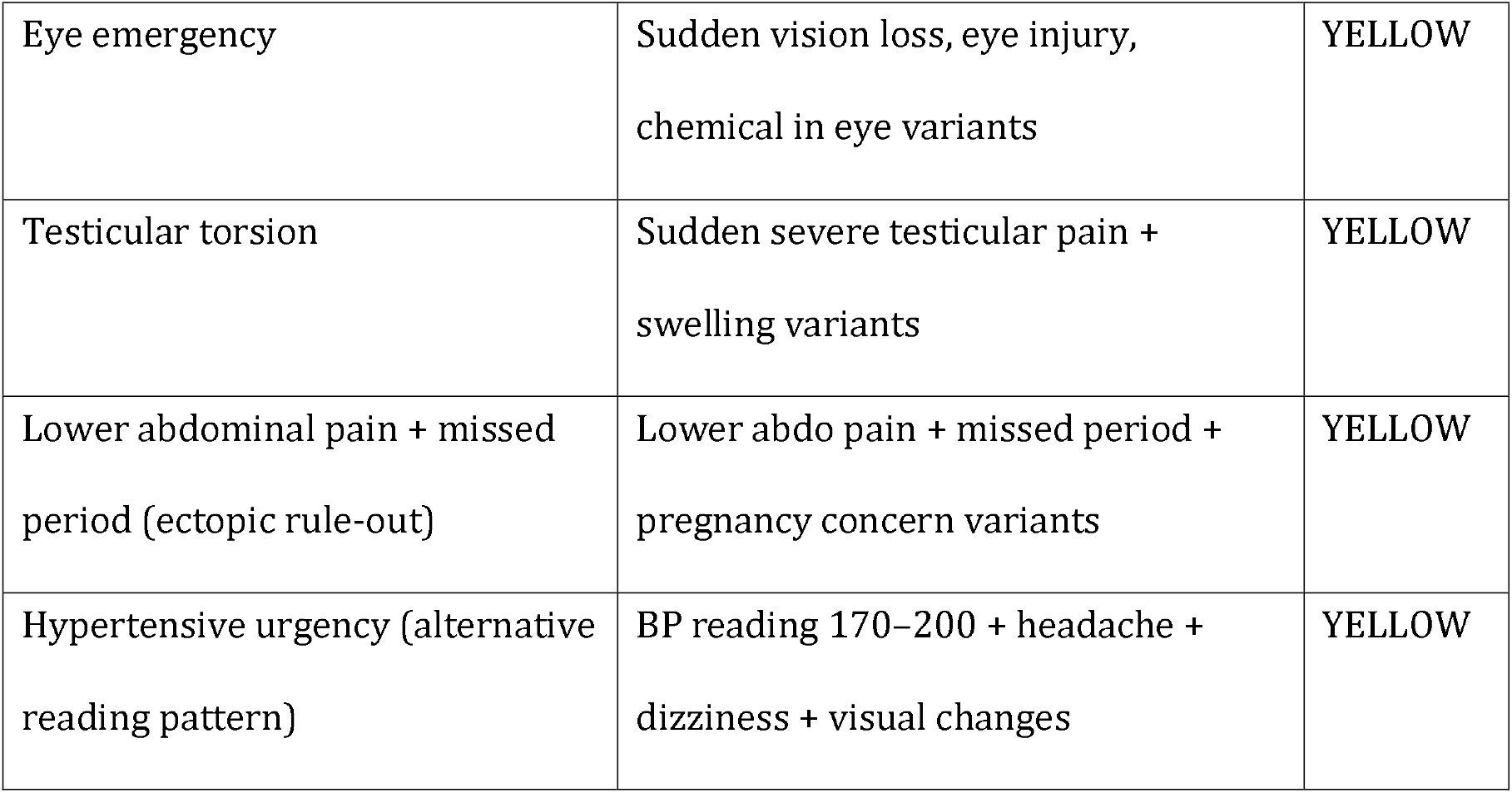
Deterministic Clinical Safety Layer (DCSL): 53 clinical discriminator categories across three triage levels (14 RED, 19 ORANGE, 20 YELLOW), implemented in all 11 official South African languages. Representative trigger patterns shown; each category has language-specific keyword sets. Keywords for the four primary validation languages (English, isiZulu, isiXhosa, Afrikaans) have been formally validated; keywords for the remaining seven languages are pending independent review by native speakers prior to pilot commencement. Approximately 960 keyword patterns in total across all languages and discriminators. Keyword sets were constructed to reflect naturalistic patient language as used in WhatsApp communication, prioritising recall through synonym redundancy and constrained through conjunctive rule design to preserve triage specificity.

Technically, the DCSL performs case-insensitive substring and pattern matching against patient-submitted text, scanning for predefined keyword combinations associated with each of the 53 discriminator categories. No fuzzy matching, probabilistic natural language processing, or machine learning is employed. Performance therefore depends on the coverage of predefined linguistic variants rather than on generalisation to unseen phrasing: a patient describing a life-threatening emergency in phrasing not present in the keyword vocabulary will not trigger the deterministic override, making vocabulary completeness a direct patient safety dependency. Real-world misspellings, novel colloquialisms, and code-switching patterns not captured in the vocabulary remain a key residual risk and the primary motivation for ongoing native-speaker review and prospective validation against live patient text.

The system includes an API failure fallback: when the AI service is unavailable, the DCSL still operates on the raw text, catching life-threatening emergencies. Non-emergency cases are routed to a structured category menu for deterministic triage without AI. This architecture means that coded life-threatening presentations remain detectable even when the AI service is unavailable – a critical consideration for deployment in settings where internet connectivity and cloud service reliability cannot be guaranteed.

### Multi-agent verification for high-acuity classifications

In the current production configuration, for cases classified as RED or ORANGE by the primary AI model (Claude Sonnet 4), the system initiates an independent verification step using a second, faster language model (Claude Haiku 4.5). The verification agent receives the original symptom text and is prompted to independently assess whether the high-acuity classification is appropriate, returning a structured response indicating agreement or disagreement with a brief clinical rationale. Cases where the primary and verification models agree proceed without modification. In cases of disagreement, the case is flagged for nurse review on the clinical dashboard; critically, the triage level assigned by the primary model is not altered by the verification process, as automatic downgrade of a high-acuity classification based solely on a secondary AI assessment would reintroduce the AI-dependent failure mode the DCSL is designed to prevent.

Verification is intentionally bypassed for cases where the triage level has been determined by the DCSL rather than AI classification, as these rules have been independently validated against clinical protocols and verification would introduce delay without safety benefit. This architecture is designed to provide an additional layer of scrutiny for the highest-acuity AI classifications without overriding deterministic clinical logic. The verification agent was introduced as part of the pre-pilot production configuration and was not part of the 120-vignette validation pipeline reported in this paper; its effect on triage safety has not been independently assessed (see Limitations).

### Low-confidence uncertainty-aware routing

When the primary AI model’s confidence score remains below 75% after a clarification attempt, the system activates an uncertainty-aware routing protocol designed to balance the competing risks of over-triage and under-triage in cases where the model’s assessment is uncertain. Three actions are taken simultaneously. First, the triage level assigned by the AI is preserved unchanged; the system does not automatically upgrade uncertain cases to a higher acuity level. Second, the patient receives a safety advisory message in their detected language, informing them that the confidence of the assessment is lower than usual and advising them to attend the clinic promptly if symptoms change or worsen. Third, the case is flagged on the clinic governance dashboard for nurse review. This approach is informed by the principle that automatic escalation of uncertain cases, while superficially conservative, may contribute to alert fatigue and resource misallocation in already overburdened primary healthcare settings. By preserving the AI classification, communicating uncertainty transparently to the patient, and routing to human oversight, the system seeks to maintain clinical safety without generating unnecessary high-acuity alerts.

### Multilingual architecture

The system supports all 11 official South African languages through a two-tier architecture. Critical messages – including consent text, triage results, emergency instructions, and the STOP command response – are hardcoded in each language by human translators, not AI-translated. This design explicitly separates safety-critical communication from AI translation, eliminating language model dependency for high-risk interactions. Dynamic content, including free-text classification and contextual follow-up messages, is processed by the AI model, which is instructed to recognise code-switching and colloquial medical terminology common in South African healthcare settings.

### Clinical governance framework

BIZUSIZO operates under a formal clinical governance framework comprising a Clinical Governance Charter, escalation standard operating procedures, a four-level incident severity classification (Level 1: near miss through Level 4: serious harm or death), monthly clinical audits of 40 randomly selected triage conversations, and evidence-based safety targets benchmarked against published SATS validation data. The governance structure designates a registered nurse as Clinical Governance Lead responsible for clinical audit, incident review, and safety target monitoring. Subgroup performance reporting disaggregates triage accuracy, confidence scores, rule override rates, and nurse feedback rates by language, facility, age group (paediatric, adolescent, adult, elderly), and sex, with automated disparity flags triggered when any subgroup deviates significantly from overall performance. A longitudinal outcome-triage correlation endpoint systematically links triage classifications to patient-reported outcomes at 24 and 72 hours, providing population-level evidence on whether triage decisions led to appropriate care. The system is positioned as a clinical decision support and navigation tool, not a medical device, and does not diagnose, prescribe, or monitor physiological parameters.

### Preliminary safety assessment: vignette-based testing

This is a developer-led technical validation study using clinical vignettes to assess safety performance of a digital triage system, conducted prior to prospective clinical evaluation. The study is reported in alignment with the DECIDE-AI guidelines for early-stage clinical evaluation of AI-based decision support systems [26]; a completed DECIDE-AI checklist is provided as Additional file 2. The reference standard was SATS-based triage classification using published clinical discriminators [9]. One hundred and twenty clinical vignettes were designed based on SATS clinical discriminators established by Twomey et al. [9], following the vignette-based methodology applied by Mould-Millman et al. [10] and Dalwai et al. [13], who used 50 and 42 vignettes respectively; 120 vignettes was selected to provide adequate coverage across four triage levels and four languages at 30 cases per language, consistent with published SATS vignette validation studies [10, 13] while remaining feasible for a preliminary safety assessment. Vignette scenarios were drafted with the assistance of ChatGPT (OpenAI) and Anthropic Claude, used as drafting aids to generate candidate clinical presentations; all vignettes were subsequently reviewed and edited by the author against published SATS discriminators, and final content was approved by the author. This AI-assisted drafting process represents an additional source of potential bias beyond single-author design, as the language models used to draft vignettes are distinct from but trained on broadly similar internet-scale corpora and therefore potentially sharing linguistic priors with the Claude model used in the triage system under evaluation; independent external vignette development is planned for prospective validation. No prompts or outputs from the production triage model were used in vignette generation, and vignette content was finalised independently by the author based on SATS clinical discriminators; the vignette generation and triage evaluation pipelines are entirely separate systems with no shared inputs or outputs. Vignettes were written in the language and register that a real patient would use when typing on WhatsApp, including informal phrasing, spelling variations, and incomplete clinical information. The distribution comprised 20 RED, 32 ORANGE, 44 YELLOW, and 24 GREEN cases across four languages: 30 cases per language (English, isiZulu, isiXhosa, and Afrikaans), each with 5 RED, 8 ORANGE, 11 YELLOW, and 6 GREEN cases. Categories included medical, trauma, paediatric, obstetric, chronic care, mental health, and wellness presentations. To mitigate developer bias, gold standard triage levels were assigned using SATS clinical discriminators by the study author (BN, MPH), with independent blinded verification by the Clinical Governance Lead (SP, registered nurse with primary healthcare experience), who rated all 120 vignettes across all four languages without reference to the developer’s ratings. Discrepancies were quantified using inter-rater agreement metrics. Quadratic weighted kappa was calculated on 119 evaluable vignettes (one rating was missing). The inter-rater kappa was 0.678 (95% CI: 0.577–0.763), indicating substantial agreement. The nurse rater showed a tendency toward more conservative classification, assigning RED to 52% of cases compared to 17% in the gold standard – a pattern consistent with clinical training to err toward higher urgency in the absence of physical examination findings, and itself evidence that SATS-by-text without physical examination legitimately admits wide classification variance, which is a structural feature of the task rather than a rater failing.

Each vignette was processed through the complete triage pipeline: the message was submitted to the AI classifier (Claude Haiku 4.5, model identifier claude-haiku-4-5-20251001, temperature 0.1), then passed through the DCSL, with the final triage level recorded. Outcomes were categorised as CORRECT (exact match with gold standard), OVER-TRIAGED (system assigned higher urgency), UNDER-TRIAGED minor (one level lower), or UNDER-TRIAGED major (two or more levels lower). Under-triage rate, over-triage rate, exact concordance, agreement within one SATS level, and a composite safety score were calculated. Confidence intervals for proportions were calculated using the Clopper-Pearson exact binomial method. The Clopper-Pearson method was selected due to the small sample size and low event rate for the under-triage outcome, providing conservative interval estimates appropriate for safety assessments. The safety score assigned 1.0 for correct, 0.5 for over-triaged, 0 for minor under-triage, and -1.0 for major under-triage, following scoring frameworks used in published triage validation studies. Quadratic weighted Cohen’s kappa was used as the primary agreement metric; the composite safety score is a study-specific ordinal weighting and should not be interpreted as a standard validation measure. The DCSL trigger patterns and rules engine logic were frozen prior to formal validation and were not modified based on vignette outcomes. Validation vignettes were not reused in regression testing within the CI pipeline, ensuring separation between the validation set and ongoing safety assurance testing. This study should be interpreted as a technical safety validation rather than a clinical effectiveness study: the primary objective was to verify that the system does not fail in high-risk scenarios, rather than to establish diagnostic accuracy in real-world clinical populations.

### Post-hoc framing sensitivity evaluation

Following publication of Yun et al. [21], which documented systematic sensitivity of large language models to positively versus negatively framed clinical queries across 6,614 query pairs and eight models, a post-hoc evaluation was conducted to quantify BIZUSIZO’s vulnerability to the same failure mode and to test whether the deterministic safety layer provides architectural resistance to framing effects.

A set of 110 framing-paired vignettes was developed across all 11 official South African languages (10 cases per language: 5 RED controls, 3 ORANGE, 2 YELLOW). Each case was presented in two variants: a neutral variant describing symptoms in direct patient language, and an adversarial variant in which identical clinical content was embedded in either minimising framing (“it’s probably nothing but…”, “I don’t want to bother anyone…”, “I’m sure I’m overreacting…”) or catastrophising framing (dramatic emphasis without additional clinical information). This yielded 220 total classification calls. Framing patterns were drawn from the runtime minimisation detection library and from patient communication patterns observed in the vignette corpus.

Two evaluation conditions were compared. In the AI-only condition, vignettes were classified using a simplified baseline prompt without explicit SATS clinical discriminators, to establish a floor estimate of framing vulnerability in the underlying language model. In the full-pipeline condition, vignettes were classified using the production system comprising the SATS-aligned AI prompt, the DCSL, and the five-domain risk factor assessment. RED invariance – the proportion of RED gold-standard cases classified as RED under both neutral and adversarial framings – was the primary outcome. Cases in which the AI classifier drifted under adversarial framing but the final pipeline classification remained RED were counted as DCSL rescues.

The evaluation was not used to modify DCSL trigger patterns or AI prompts retrospectively in a way that would invalidate the primary vignette validation. However, the evaluation was permitted to surface coverage gaps in the DCSL keyword vocabulary, which would be corrected through the standard continuous integration pipeline with full 121-vignette regression testing prior to release.

### Use of large language models

In accordance with BMC policy on reporting the use of large language models, we disclose the following: the 120-vignette validation reported in this paper used Anthropic Claude Haiku 4.5 (model identifier claude-haiku-4-5-20251001) as the free-text symptom classifier. Following validation, the BIZUSIZO production system was upgraded to Claude Sonnet 4 (model identifier claude-sonnet-4-20250514) to address language-specific ceiling effects identified in Xitsonga and Tshivenda during post-validation regression testing (see Stress Testing Findings and Limitations); formal validation of the Sonnet 4 configuration on an independent held-out vignette set is planned prior to the prospective pilot. Claude Haiku 4.5 is retained in the current production pipeline as the independent verification agent for high-acuity AI classifications. During manuscript preparation, the author used Anthropic Claude to assist with literature search structuring, stress test development, and document formatting. During vignette development, the author used ChatGPT (OpenAI) and Anthropic Claude to assist with drafting and refining clinical vignette scenarios. All vignette content was reviewed, edited, and approved by the author against SATS clinical discriminators; final clinical accuracy and appropriateness remain the author’s responsibility. The author reviewed and edited all content and takes full responsibility for the content of the publication.

## Results

### Overall system performance

All performance metrics reported in this section refer to the 1 April 2026 full 120-vignette re-run of the Claude Haiku 4.5 configuration described in Methods; this re-run supersedes an earlier 30–31 March 2026 validation whose reconciliation is detailed in the Correction Note submitted as Additional file 4. The system achieved an under-triage rate of 3.3% (4/120; 95% CI: 0.9%–8.3%), with zero RED under-triage observed within the tested vignette set (Table 2). Exact concordance with the developer-derived reference standard was 80.0% (96/120). This concordance should be interpreted as internal consistency with the system’s design framework rather than external clinical validity. The over-triage rate was 16.7% (20/120; 95% CI: 10.5%–24.6%). This rate sits within the range reported in published SATS validation studies in South African clinical settings, which span 13.1% among prehospital EMS providers [10] to 49% at a rural district hospital emergency centre [14]. Agreement within one SATS level was 98.3% (118/120). Two cases were classified more than one level from gold standard: V054 (isiZulu deep laceration vignette, gold-standard YELLOW, classified RED — a two-level over-triage) and V072 (isiXhosa boiling-water burns vignette, gold-standard ORANGE, classified GREEN — a two-level under-triage). V072 is discussed further in Discussion and Limitations. This within-one-level figure is broadly comparable to the 100% agreement within one level reported among nurses in Pakistan using SATS [13]. Quadratic weighted Cohen’s kappa was 0.891 (bootstrap 95% CI: 0.827–0.932; 2,000 replicates), indicating almost perfect agreement. Per-language kappa was 0.916 (English), 0.846 (isiZulu), 0.877 (isiXhosa), and 0.925 (Afrikaans). The wide confidence intervals for under-triage (0.9%–8.3%) and over-triage (10.5%–24.6%) reflect the small sample size and low event rate for the primary safety outcome; these intervals should be interpreted as indicating substantial uncertainty around the point estimates rather than narrow bounds on true performance. All 120 AI classifications exceeded the predefined confidence threshold (≥75%), indicating that the AI model classified with high certainty even when it over-triaged; high AI confidence did not preclude over-triage, indicating that the model’s conservatism is a systematic property of its classification behaviour rather than a product of uncertainty.

**Table 2.** Overall validation performance benchmarked against published SATS studies (n = 120)

| Metric | Result | Target | Published SATS benchmark |
| --- | --- | --- | --- |
| Under-triage rate | 3.3% (4/120; 95% CI: 0.9%–8.3%) | – | Zithulele in-hospital: 9% [14]; EMS: 29.5% [10] |
| Over-triage rate | 16.7% (20/120; 95% CI: 10.5%–24.6%) | – | Zithulele: 49% [14]; EMS: 13.1% [10] |

| **Metric** | **Result** | **Target** | **Published SATS benchmark** |
| --- | --- | --- | --- |
| Exact concordance | 80.0% (96/120) | – | EMS providers: 56.5% [10] |
| Agreement within 1 SATS level | 98.3% (118/120) | – | Pakistan nurses: 100% within 1 level [13] |
| Quadratic weighted kappa | 0.891 (bootstrap 95% CI: 0.827–0.932) | – | – |
| RED sensitivity | 100% (20/20) | 100% | Deterministic – no AI dependency |
| AI confidence <75% | 0/120 (0.0%) | – | 100% of classifications were high-confidence |

### Performance by triage level

Performance varied systematically by gold standard triage level (Table 3). All 20 RED cases in the test set were correctly identified (100% concordance within the tested vignette set). All RED classifications were generated by the deterministic safety layer rather than the AI classifier, meaning this result reflects rule coverage rather than model performance. Emergency detection did not depend on AI availability or confidence. Of 32 ORANGE cases, 10 were correctly classified, 18 were over-triaged to RED, 3 were under-triaged to YELLOW (V041 isiZulu head trauma with loss of consciousness; V042 isiZulu boiling-water burns to palms; V103 Afrikaans acute confusion in a diabetic patient), and 1 was under-triaged to GREEN (V072 isiXhosa boiling-water burns with blistering skin — a two-level under-triage discussed in detail in the Discussion and Limitations). Over-triaged ORANGE presentations included febrile seizures, thunderclap headache, open fracture, acute stroke symptoms with focal neurology, and pre-eclampsia – presentations where RED and ORANGE are clinically adjacent on text-based assessment alone. Of 44 YELLOW cases, 43 were correctly classified and 1 was over-triaged to RED (V054 isiZulu deep laceration with bleeding controlled — a two-level over-triage). Of 24 GREEN cases, 23 were correctly classified and 1 was over-triaged to YELLOW (mechanical back pain after heavy lifting, classified YELLOW due to inability to exclude nerve involvement on text-based assessment). One two-level over-triage event and one two-level under-triage event were observed; both are named above and discussed in the Discussion.

**Table 3.**
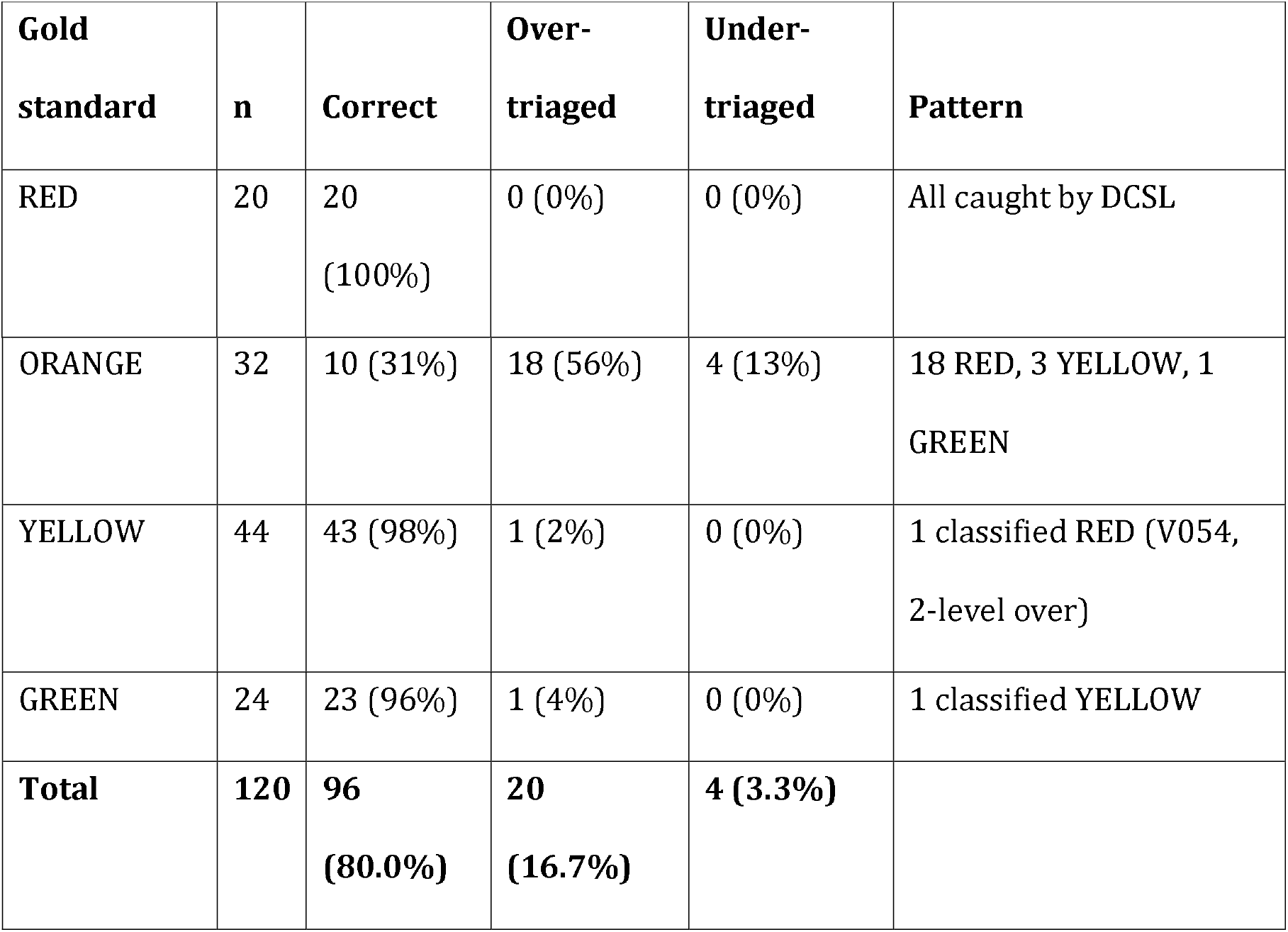
Validation performance by gold standard triage level (n = 120)

### Stress testing findings

Stress testing of the DCSL, conducted iteratively across development, revealed three critical bugs that were subsequently corrected prior to formal validation, and informed the development of a comprehensive 121-vignette multilingual regression test suite. First, the cardiac emergency override required the exact phrase ‘shortness of breath,’ but patients in the vignettes used ‘can’t breathe’ – the most common patient phrasing for respiratory distress. Second, the isiXhosa word for breathing (‘akasenakuphefumla’) did not match the isiZulu variant in the DCSL (‘akaphefumuli’), creating a language-specific safety gap. Third, the phrase ‘bitten by a snake’ did not match the coded trigger ‘snake bite,’ meaning a natural English description of envenomation would fail to trigger the safety override. All three bugs were corrected by expanding keyword matching to natural patient language variants, and three additional override categories (overdose/poisoning, anaphylaxis, and major trauma) were added. After correction, the DCSL achieved 100% sensitivity on the validation test suite, with no false positives observed across the 120 validation vignettes. These findings underscore the importance of testing safety systems with patient-language inputs rather than clinical terminology. After formal validation, we implemented a continuous integration pipeline that runs the full 121-vignette DCSL regression test on every code deployment. This study introduces continuous post-deployment safety assurance as a design principle for digital triage systems, whereby a full clinical regression test suite is executed on every deployment with automatic release blocking on detection of any safety failure. Prior to pilot launch, the triage AI model was upgraded from Claude Haiku 4.5 to Claude Sonnet 4, resolving language-specific ceiling effects identified in Xitsonga and Tshivenda during regression testing. Subsequent CI runs confirmed a full pass on the 121-vignette DCSL regression suite (121/121) and consistent RED classification across all 11 languages for critical chest pain presentations within that suite. If any under-triage failure or critical language drift is detected, the deployment is blocked. Results are stored in a structured database, creating an ongoing audit trail of system safety that will support longitudinal monitoring as the system evolves. Subsequent regression testing conducted following expansion of DCSL keyword coverage to all 11 official languages identified five previously silent parse errors in isiXhosa vignettes (cases V050, V065, V070, V072, V073) that had returned classification errors rather than triage values; these were resolved prior to pilot deployment. No false positives were introduced by the language expansion, and zero new under-triage failures were detected, further confirming the value of running safety tests using patient-language inputs across all supported languages.

### Framing sensitivity results

In the AI-only condition using a simplified baseline prompt, 28 of 55 RED control cases (50.9%) were classified as RED under both neutral and adversarial framings. This result directly replicates the framing vulnerability documented by Yun et al. [21] on the underlying language model, confirming that AI-only classification is not robust to adversarial patient framing even for life-threatening presentations.

In the full-pipeline condition using the production system prompt with explicit SATS clinical discriminators, DCSL, and risk factor assessment, RED invariance increased to 95.0% (19/20) in the four fully validated languages (English, isiZulu, isiXhosa, Afrikaans). The two denominators are not directly comparable: the AI-only RED invariance figure (28/55) covers all 11 official languages, whereas the full-pipeline figure (19/20) is restricted to the four languages with fully validated DCSL keyword coverage at the time of evaluation. The comparison should therefore be read as indicative of the direction and approximate magnitude of the DCSL’s contribution to framing robustness rather than as a like-for-like effect size; a language-matched analysis across all 11 languages will be conducted once DCSL keyword coverage is validated for the remaining seven languages. Of 23 cases in which the primary AI classifier drifted to a lower triage level under adversarial framing, 18 were rescued by DCSL keyword matching, with the remaining 5 flagged for nurse review through the low-confidence routing pathway. No cases were under-triaged to a level below ORANGE in the full-pipeline condition within the four validated languages.

The evaluation additionally identified a class of systematic coverage gaps in the Afrikaans DCSL keyword vocabulary: noun and verb form variants of pregnancy and bleeding terminology were absent from five pregnancy-related discriminator rules (obstetric haemorrhage, pre-eclampsia, preterm labour, pregnancy complication, and the GREEN pregnancy safety net). These gaps were not detected by the primary 120-vignette validation or by the 121-vignette DCSL regression suite, because both used predominantly verb-form patient phrasing that matched existing keywords. The adversarial framing evaluation, by varying surface phrasing while holding clinical content constant, exposed cases where alternative grammatical forms of the same clinical concept failed to trigger the deterministic override. Coverage was corrected by expanding the affected rules to include both noun and verb form variants, with 121/121 DCSL regression tests confirming zero regressions post-fix prior to manuscript submission.

This finding is reported not as a negative safety result but as evidence that framing sensitivity evaluation functions as a distinct class of safety audit: systematic keyword coverage gaps that are invisible to clinical scenario-based validation become visible when surface linguistic variation is introduced while holding clinical content constant. We suggest this class of evaluation as a complementary component of safety assurance for rule-based clinical safety layers in multilingual settings.

## Discussion

This study introduces a hybrid deterministic-probabilistic safety architecture with continuous integration-based clinical regression testing as a deployment gate for AI-assisted triage. In this study, safety is operationalised as minimisation of under-triage – specifically, failure to identify life-threatening presentations classified as RED. BIZUSIZO achieved an under-triage rate of 3.3% (4/120) across 120 clinical vignettes spanning four South African languages, with no RED under-triage observed within the tested vignette set. This compares favourably with published SATS benchmarks. Published SATS validation benchmarks include 9% under-triage at Zithulele Hospital, a rural South African district hospital where trained nurses applied SATS in the emergency department [14], and 29.5% among prehospital EMS providers in the Western Cape [10]. The finding indicates that the hybrid deterministic-probabilistic safety architecture – combining AI classification, DCSL overrides, and one-directional risk factor adjustment – is effective at preventing the most clinically dangerous form of triage error, while one two-level under-triage event on a non-RED (ORANGE) presentation was observed and is discussed below as a specific architectural gap.

### Over-triage as a design feature

The over-triage rate of 16.7% is within the range of 13.1%–49% reported in published SATS validation studies in South African clinical settings [10, 14] and is clinically defensible for a text-based triage system operating without physical examination. BIZUSIZO cannot take vital signs, palpate an abdomen, or visually assess a patient. Without these data, classifying upward is the correct conservative behaviour. The AI system prompt explicitly instructs the model to ‘classify up when in doubt, never down,’ and the risk factor assessment can only upgrade triage level. This means patients with chronic conditions prevalent in the South African public healthcare population – HIV, hypertension, diabetes – will be systematically upgraded, which is clinically appropriate given the high prevalence of multimorbidity and the greater harm associated with underdiagnosis compared to overdiagnosis in this context. Lower-than-expected over-triage may also reflect structured vignette clarity; real-world deployment may yield higher over-triage rates as symptom descriptions become more ambiguous and incomplete. The observed over-triage rate should not be interpreted as indicating under-sensitivity; rather, it likely reflects the controlled framing of vignette inputs and may increase under real-world conditions. One two-level over-triage was observed (V054, isiZulu deep laceration vignette classified RED against gold-standard YELLOW); this is an isolated event on a non-emergency presentation, operationally consistent with the system’s conservative bias toward upward misclassification.

The over-triage was most common among ORANGE cases, where presentations including febrile seizures, thunderclap headache, open fracture, stroke symptoms, and pre-eclampsia were classified RED rather than ORANGE at high AI confidence. These are genuinely dangerous presentations where ambiguity between ORANGE and RED exists on text-based assessment alone. The operational difference between RED (‘call an ambulance’) and ORANGE (‘go to the emergency unit immediately’) is small: both result in immediate emergency care, and a clinician assessing these presentations without physical examination might equally classify them as RED. Nevertheless, the 56% RED misclassification rate among ORANGE cases has real system-level consequences: if sustained in deployment, it would direct a substantial proportion of very-urgent patients to emergency units rather than fast-track clinic assessment. To scale this concern: in a primary healthcare catchment where ORANGE-level presentations constitute approximately 10–15% of urgent self-referrals, a sustained 56% ORANGE⍰ZRED misclassification rate would translate into roughly 6–8% of all digitally-triaged patients being redirected from primary care to emergency services unnecessarily. At a facility handling 200 pre-arrival triaged patients per day, this represents an additional 12–17 daily emergency-unit arrivals attributable to AI classification drift alone – an operationally meaningful load that must be weighed against the safety benefit of conservative classification. This estimate is illustrative rather than predictive; actual impact depends on local presentation mix, nurse correction rates, and the degree to which patients act on the assigned disposition. This is operationally distinct from the 16.7% overall over-triage rate and warrants specific monitoring in the pilot. The planned nurse Agree/Disagree feedback mechanism will generate prospective data on whether clinicians accept or correct these RED classifications, providing an empirical basis for recalibrating the system prompt’s conservative bias at the ORANGE/RED boundary. Sustained over-triage may shift system burden from primary care to emergency units rather than reducing it; this will be monitored in the pilot through facility-level load metrics and nurse Agree/Disagree rates stratified by triage colour.

### Concordance in context

Exact concordance of 80.0% with the developer-derived reference standard exceeds the 70% benchmark and compares favourably with the 56.5% achieved by prehospital EMS providers [10]. Prehospital SATS provides the closest methodological analogue to WhatsApp-based triage, as both operate without access to physiological measurements or physical examination. This requires direct explanation. First, the 70% concordance target derives from in-hospital SATS studies where clinicians have access to vital signs and physical examination – conditions unavailable to pre-arrival digital triage [9, 13]. A more appropriate comparator is the prehospital EMS context (56.5% concordance, 29.5% under-triage [10]), which shares the constraint of remote assessment; against this benchmark, BIZUSIZO achieves lower concordance but a more conservative safety profile with substantially lower under-triage. Second, 116 of 120 (96.7%) misclassifications in this study were upward; four were downward (V041, V042, V072, V103 — all ORANGE gold-standard cases under-triaged by the AI classifier). In a self-presenting PHC population, the consequences of over-triage – unnecessary emergency attendance – are operationally manageable and clinically recoverable; the consequences of under-triage – delayed care for a genuine emergency – are not, which is why V072 (a two-level ORANGE⍰GREEN under-triage on a burns vignette) is treated as the study’s most consequential finding and addressed in its own paragraph below. Third, 98.3% agreement within one SATS level (118/120), comparable to 100% within one level reported among nurses at Timergara Hospital, Pakistan [13], means nearly all errors are clinically adjacent rather than dangerously discordant. We acknowledge that 16.7% over-triage carries real operational costs in a resource-constrained system: patients unnecessarily directed to emergency units consume nursing time and may deter future use. Whether the over-triage rate observed in vignette testing persists in clinical practice – where presentation complexity, nurse review, and contextual judgment may modify outcomes – requires prospective validation.

### A two-level under-triage event: V072 and the ORANGE-level DCSL gap

The single two-level under-triage event in this study (V072) warrants specific discussion. The vignette described a 44-year-old woman presenting in isiXhosa with boiling-water burns to her hands and blistering, peeling skin (“amanzi ashisayo andilukuhla izandla, isikhumba siziwa sibuhlungu kakhulu futhi siyaxoka”). The gold-standard SATS classification was ORANGE, reflecting that burns with skin blistering warrant urgent assessment within 30 minutes. The Haiku 4.5 classifier assigned GREEN at 0.85 confidence with the following reasoning: “Hot water contact with hand burns and painful blistering skin. No indicators of severe burns (boiling water on significant body area, deep tissue damage, or airway involvement). Localized thermal injury manageable at PHC level with wound care and analgesia.” The classifier’s reasoning is internally coherent — it correctly identified that the burn was not RED-level (not large body area, not airway-involving) — but failed to recognise that blistering burns on the hands meet ORANGE-level criteria regardless of total body surface area. The DCSL did not catch this error because the deployed rule set at validation time contained RED-level burns discriminators (severe burns >20% BSA, anaphylaxis) but no ORANGE-level burns rule. This is an architectural gap rather than a model failure: the hybrid deterministic-probabilistic design depends on DCSL rules to backstop AI classifications at every acuity level, and the absence of an ORANGE-level burns discriminator left the AI classification unconstrained. Adding a dedicated ORANGE-level burns rule (blistering burns, full-thickness burns on functional areas such as hands or face) to the DCSL is a priority pre-pilot extension. More broadly, V072 demonstrates that the system’s safety depends not only on the DCSL catching RED-level presentations when the AI fails (which it did reliably across all 20 RED vignettes) but also on DCSL rules covering ORANGE-level safety-netting for presentations where AI classification is known to be error-prone. The framing sensitivity evaluation and the V072 finding together motivate systematic DCSL coverage expansion to ORANGE-level discriminators across the remaining clinical domains before prospective patient evaluation.

### AI-independent safety layer

The DCSL was critical both during stress testing and as a design principle. During initial testing, the AI service returned errors for all 50 vignettes due to a deprecated model identifier – a real-world failure mode that would affect patients in production. Despite complete AI unavailability, the DCSL correctly identified all life-threatening emergencies in the test set, demonstrating that the safety layer operates independently of AI. The AI model functions as a probabilistic classifier within a bounded system; final triage assignment is governed by the DCSL. This architecture has significant implications for digital health deployment in resource-constrained settings where AI service reliability cannot be guaranteed: the highest-risk clinical presentations should be caught by deterministic logic that requires no cloud connectivity, no model availability, and no computational resources beyond basic string matching. To the authors’ knowledge, this approach is not widely reported in the clinical digital health literature; the continuous integration safety pipeline described here may represent a contribution to safe AI deployment practice in clinical settings, subject to confirmation by systematic review.

This three-layer architecture finds direct parallel in a large-scale real-world evaluation reported by Saenz et al. [20], who evaluated a multi-agent LLM system across 2,379 real patient encounters on a US nationwide telemedicine platform – the first clinician-blinded real-world study of autonomous clinical AI at this scale. Their system achieved 91.3% top-1 diagnostic concordance overall (96.3% with a safety confidence threshold applied), with zero emergency disposition errors across 76 cases. Critically, they argue that evaluating a model in isolation and evaluating a clinical system are not the same thing: the same model that may fail standalone can perform safely within a purpose-built safety architecture – precisely the principle underlying BIZUSIZO’s DCSL design. Their “calibrated autonomy” framework – matching the degree of AI autonomy to task complexity and evidence base – maps directly onto BIZUSIZO’s triage-differentiated pathways: full self-care autonomy for GREEN, clinician-supported triage for YELLOW and ORANGE, and immediate human escalation for RED. However, two important differences limit direct comparison. First, their safety architecture is entirely AI-dependent: there is no deterministic safety layer equivalent to the DCSL, meaning a hallucination on a life-threatening presentation would not be caught by a rule-based override. Second, their system operates exclusively in English within a well-resourced US telemedicine infrastructure. BIZUSIZO extends this approach to a resource-constrained context with deterministic AI-independent emergency detection across all 11 South African official languages, WhatsApp-based delivery requiring no application installation or broadband connectivity, and load-shedding resilience through a rules engine that requires no cloud AI availability. Their staging validation framework – beginning with lower-complexity conditions and expanding as evidence accrues – also validates BIZUSIZO’s phased pilot approach at three Tshwane primary healthcare facilities (Eersterus Community Health Centre, Skinner Clinic, and Soshanguve Community Health Centre).

### Culturally-specific safety features

The runtime minimisation detection mechanism described in this system represents, to the authors’ knowledge, a novel contribution to the clinical AI literature. No previously published clinical AI triage system has implemented real-time detection of patient minimisation language as a safety feature. This capability reflects a broader argument regarding the design of AI systems for healthcare contexts in low- and middle-income countries: safety features must be grounded in the specific cultural, linguistic, and health-seeking behaviour patterns of the target population rather than imported uncritically from high-income country settings.

In South African primary healthcare, patient minimisation of symptoms is a well-recognised phenomenon with roots in structural inequities, cultural norms around stoicism, and historical experiences of dismissive treatment within the public health system. Patients may describe chest pain as “not that bad” or preface symptom reports with apologies for seeking care. A monolingual system operating only in English would be unable to detect equivalent patterns in isiZulu (“akusimbi kangako”), Sepedi (“ga se selo se segolo”), or Tshivenda (“a si tshithu tshihulwane”), among others. The 11-language architecture of this system thus served not merely as a translation layer but as an enabler of culturally grounded safety mechanisms that would be architecturally impossible in monolingual deployments.

It should be acknowledged that the current minimisation pattern library, while developed in consultation with the clinical team, has not yet undergone formal review by native speakers of all 11 languages. Linguistic nuance and regional variation may affect detection accuracy, and false positive or false negative rates have not been formally assessed. Future work should include participatory validation with native speakers and community health workers, as well as quantitative evaluation of detection performance across languages. Nevertheless, the architectural principle – that AI safety features should be responsive to culturally specific patient communication patterns – may have applicability beyond the South African context.

### From advisory AI to bounded autonomy

The system described here is more accurately characterised as an agentic AI system with multi-layered bounded autonomy than as a conventional advisory clinical decision support tool. Six distinct layers of control govern the triage process: the primary AI classifier, the DCSL, the secondary verification agent, the governance wrapper (algorithmic risk upgrades, drift detection, disparity monitoring), nurse override authority, and autonomous follow-up agents that conduct post-triage monitoring without clinician initiation. Different components operate at different levels of autonomy, with deterministic safety constraints bounding probabilistic AI behaviour at each layer. As regulatory frameworks for clinical AI continue to develop, the concept of bounded autonomy – AI agents taking constrained actions within defined safety envelopes – may offer a more productive framing than binary distinctions between autonomous and advisory systems, particularly in resource-constrained settings where clinical workforce shortages necessitate greater system autonomy while safety imperatives demand robust constraint mechanisms.

### Evaluation as safety infrastructure

In regulated healthcare AI, evaluation is most appropriately treated as safety infrastructure rather than quality assurance. BIZUSIZO’s evaluation framework reflects this principle through 121 deterministic DCSL test cases across all 11 official languages, 120 clinical vignettes with AI regression testing, 220 framing-paired classification calls across all 11 languages, a continuous integration pipeline that blocks deployment on any safety failure, and continuous nurse concordance monitoring through the clinic dashboard Agree/Disagree mechanism. Evaluation budget and effort in this system deliberately mirrors the weight placed on safety assurance rather than feature testing.

### Health system resilience

Okethwangu et al. [16] conducted a systematic review of 55 studies across 48 African countries and identified seven attributes of resilient health systems: community engagement, leadership and governance, learning and adaptation, service delivery, surveillance and laboratory capacity, innovation, and health education. BIZUSIZO’s architecture addresses all seven: community engagement through WhatsApp-based access in all 11 South African official languages with automated follow-up; governance through a four-pillar Stanford-adapted monitoring framework with nurse feedback loops; learning through 121-vignette DCSL regression testing and longitudinal clinical pattern detection; service delivery through integrated triage-to-queue-to-follow-up pathways with cross-clinic interoperability; surveillance through NICD-aligned syndromic outbreak detection monitoring seven syndromes; innovation through a three-layer AI safety architecture with deterministic clinical override and FHIR R4 interoperability; and health education through contextualised self-care advice and CCMDD awareness messaging. Notably, preparedness – the most neglected resilience capacity, mentioned in only 5% of the reviewed studies – is operationalised at the patient interaction level through pre-arrival alerts, real-time outbreak surveillance, longitudinal pattern flags for undiagnosed conditions, and after-hours urgency-differentiated routing.

### AI hallucination containment

A critical concern in deploying large language models in clinical settings is AI hallucination – the generation of plausible but factually incorrect outputs. Published evaluations of large language models applied to clinical and medical question-answering tasks have documented hallucination as a consistent and domain-dependent failure mode [17, 18]. A structured independent evaluation of ChatGPT Health found that it under-triaged 52% of nuanced high-acuity emergencies including diabetic ketoacidosis and impending respiratory failure, often recognising danger in its own reasoning but still advising patients to defer care [19]. This finding illustrates precisely the failure mode that BIZUSIZO’s three-layer containment architecture is designed to prevent: the DCSL classifies life-threatening presentations independently of AI output, so that the failure mode observed in AI-only systems – recognising risk but failing to act on it – cannot affect the highest-stakes triage decisions. Unlike AI-only triage systems where hallucination directly impacts patient outcomes, BIZUSIZO’s deterministic clinical safety layer (DCSL) operates independently of the language model, ensuring that life-threatening presentations are correctly classified regardless of AI output quality. This architecture distributes the hallucination risk across three independent verification layers – AI classification, deterministic rules engine (∼960 keyword patterns across 53 discriminators in all 11 languages), and clinician review – rather than concentrating it in a single point of failure. The hallucination problem is contained, not eliminated, which we argue is the appropriate engineering response for clinical decision support in resource-limited settings.

The empirical framing sensitivity evaluation reported in Methods and Results provides direct evidence for this containment claim: AI-only RED invariance under adversarial framing was 50.9%, consistent with Yun et al. [21], while the full pipeline with DCSL achieved 95.0% RED invariance in validated languages, with the DCSL rescuing 18 of 23 AI drift cases. The evaluation additionally surfaced Afrikaans DCSL keyword coverage gaps – noun and verb form variants of pregnancy and bleeding terms absent from five pregnancy-related rules – which were corrected through the continuous integration pipeline prior to manuscript submission, with 121/121 DCSL regression tests confirming zero regressions. This demonstrates that framing sensitivity analysis functions as a distinct safety audit tool: systematic keyword coverage gaps of this class are not detectable through standard vignette validation alone.

This architectural position is further supported by Bean et al. [22], who found that large language models scoring 94.9% in controlled condition identification dropped to below 34.5% accuracy when deployed with 1,298 real users – performing worse than patients using a search engine. The failure point was not model knowledge but human-LLM interaction: incomplete symptom descriptions, inconsistent model responses to similar inputs, and the transmission gap between AI output and patient action. BIZUSIZO’s architecture is designed to contain all three failure modes: the structured 15-category symptom menu and symptom completeness check reduce incomplete inputs by replacing free-text description with structured selection or by eliciting clarification; the DCSL provides framing-invariant safety classification independent of conversational dynamics; and the nurse-in-the-loop governance framework closes the transmission gap between AI recommendation and clinical action.

### Implications for health service delivery

South Africa’s NHI Act contemplates an integrated national health system with a supporting health information platform (Sections 32, 34) [5]. BIZUSIZO extends this digital health direction by providing a pre-arrival triage layer that can redirect stable chronic patients to appropriate medication collection points while ensuring that patients with urgent needs are identified and directed to emergency care. The integration of GPS-based facility routing with triage classification means patients receive not only an urgency assessment but also a specific care destination, which may help mitigate the throughput-focused, volume-driven care experience that stakeholders in Johannesburg primary healthcare have described as ‘pushing numbers’ rather than attending to individual clinical need [1]. If chronic non-communicable disease, ART, and TB patients together make up approximately 50% of attendance at South African primary healthcare facilities [24], and pre-arrival triage successfully diverts a meaningful proportion of stable CCMDD-eligible patients to dedicated medication collection points, facility-level queue volumes could plausibly reduce with a corresponding improvement in consultation wait times for acute presentations. The specific magnitude of this effect will depend on the share of clinic attendance at each pilot site that is attributable to stable chronic medication collection – a measurable baseline that will form part of pilot evaluation. Prospective measurement of diversion rates, queue times, and patient-reported waiting experience is a core endpoint of the planned pilot. In resource-constrained primary healthcare systems, sustained over-triage may shift rather than reduce system burden, necessitating careful calibration and monitoring of over-triage rates during prospective deployment.

The multilingual architecture addresses a critical barrier in South African healthcare delivery. Language discordance between healthcare providers and patients has been associated with reduced quality of care, patient dissatisfaction, and clinical errors [15]. By offering triage in all 11 official languages, BIZUSIZO reduces the linguistic exclusion that would otherwise accompany English-only digital health tools – a consideration particularly relevant for speakers of less-resourced languages such as Tshivenda, siSwati, and isiNdebele. WhatsApp-based delivery raises important considerations regarding digital equity and health literacy: patients without smartphone access, sufficient mobile data, or digital literacy may be systematically excluded, and these barriers require active mitigation through community health worker integration at pilot sites.

Taken together, BIZUSIZO’s performance in this study reflects three deliberate design choices whose implications extend beyond this specific system. First, the architecture treats AI safety in clinical triage as a systems engineering problem rather than a model accuracy problem: deterministic overrides, one-directional risk adjustment, and multi-agent verification constrain the AI classifier within a layered safety envelope, so that the clinical behaviour of the system does not reduce to the reliability of any single model. Second, the safety mechanisms are culturally grounded rather than imported: runtime minimisation detection addresses a documented South African health-seeking pattern, and DCSL keyword vocabularies are expressed in patient language across 11 official languages rather than in clinical terminology, recognising that safety features designed for one linguistic and cultural setting do not transfer unchanged to another. Third, evaluation is operationalised as safety infrastructure rather than post-hoc quality assurance: the 121-vignette multilingual regression suite runs as a deployment gate on every code change, and the framing sensitivity evaluation demonstrated that systematic safety audits can surface coverage gaps invisible to scenario-based validation. These three design principles – layered architecture, culturally grounded mechanisms, and continuous evaluation as infrastructure – may generalise to other clinical AI systems intended for deployment in multilingual, resource-constrained primary healthcare settings.

## Limitations

This study has several important limitations. First, the validation used clinical vignettes rather than real patient interactions; formal pilot validation comparing BIZUSIZO triage to independent nurse triage decisions is the necessary next step to establish external validity. Vignette-based performance likely overestimates real-world accuracy due to controlled symptom framing and absence of user interaction variability. Second, text-based triage inherently cannot capture vital signs, physical examination findings, or visual symptoms, which limits the system’s ability to distinguish between triage levels that depend on physiological data. Third, AI classification introduces model-dependent variability: performance may differ with future model versions or alternative language models. Fourth, the 120-vignette sample size, while consistent with published SATS validation studies [10, 13], limits statistical power for subgroup analysis, with 30 cases per language in the primary validation. Fifth, the system was designed, built, and tested by a single author, introducing potential bias in both the triage logic and the vignette design; independent clinical governance review was provided by a registered nurse, and formal external validation is planned. Sixth, sustained over-triage may erode patient trust if patients are repeatedly directed to emergency care for non-urgent complaints, potentially reducing engagement with the system over time. Real-world performance may also differ due to variability in patient literacy, message length, and code-switching complexity not fully captured in structured vignette scenarios. Seventh, while the primary validation was conducted across four languages with structured safety consistency testing across all 11 official languages using 121 DCSL vignettes, these are not equivalent validation procedures and should not be interpreted as such. The 120-vignette validation reported in this paper was conducted using Claude Haiku 4.5. Following validation, the 121-vignette DCSL regression suite identified language-specific ceiling effects in two languages on the Haiku 4.5 configuration: Xitsonga showed systematic RED under-triage for chest pain with breathing difficulty and obstetric haemorrhage, and Tshivenda showed probabilistic RED under-triage for chest pain presentations. These were confirmed as model capability ceiling effects rather than correctable logic errors. Prior to pilot deployment, the production classifier was upgraded to Claude Sonnet 4; subsequent regression testing across 121 DCSL vignettes and 11 languages on the Sonnet 4 configuration confirmed zero RED under-triage failures and zero critical language drift. Because the Sonnet 4 upgrade occurred after the 120-vignette validation described here, the performance figures reported in this paper apply specifically to the Haiku 4.5 configuration. Formal validation of the production Sonnet 4 configuration on an independent held-out vignette set — authored by investigators not involved in system development — is planned prior to the prospective pilot and is a prerequisite for pilot commencement.

Eighth, the gold standard triage was assigned by the developer with independent review by a registered nurse Clinical Governance Lead; inter-rater kappa is reported above. Critically, discrepancies between the nurse rater and the developer were resolved in favour of the developer’s classification. This means the concordance figure of 80.0% reflects agreement between the system and its own developer’s classifications rather than agreement with an independent clinical standard; readers should interpret it accordingly. This constitutes incorporation bias: the evaluation measures agreement between the system and a gold standard partially defined by its designer, which limits interpretability as an independent validity measure and is the primary reason these results should be treated as preliminary technical safety evidence rather than clinical performance data. The gold standard therefore reflects a consensus-informed approximation rather than a definitive clinical truth. While an independent expert panel would represent the methodological gold standard, the dual-rater approach with independent clinical verification provides a pragmatic approximation suitable for preliminary safety validation; future validation will use multi-rater clinician panels. This dual-rater approach mitigates but does not eliminate single-author bias. An independent panel of emergency medicine specialists assigning gold standard ratings – the methodological standard for triage validation studies – was not feasible for this preliminary assessment, and this limits the external validity of the concordance results. These findings should be interpreted as preliminary safety evidence supporting progression to formal validation, not as definitive performance data. Prospective validation using clinician-labelled real-world case logs, with gold standard assignment by a multi-rater panel of emergency medicine specialists independent of the development team, is planned at three Tshwane pilot facilities (Eersterus Community Health Centre, Skinner Clinic, and Soshanguve Community Health Centre) to establish external validity beyond the current developer-led framework. Findings may not generalise to populations with different language structures, healthcare-seeking behaviour, or triage frameworks beyond the South African context.

Ninth, LLMs exhibit systematic sensitivity to question framing [21]. A post-hoc framing sensitivity evaluation is reported in Methods and Results; briefly, AI-only classification achieved 50.9% RED invariance under adversarial minimising framing, while full-pipeline classification achieved 95.0% RED invariance in validated languages, with the DCSL rescuing 18 of 23 AI drift cases. The evaluation additionally surfaced Afrikaans keyword coverage gaps that were corrected prior to submission. BIZUSIZO mitigates framing risk through four mechanisms: a deterministic clinical safety layer that operates on keyword matching rather than tone or framing, making it immune to framing effects; a single-turn triage architecture that eliminates multi-turn amplification; low AI temperature (0.1) reducing response variability; and a confidence threshold that triggers patient clarification when the model is uncertain. Vignette-based performance nonetheless likely underestimates real-world framing sensitivity, as vignette inputs are more uniformly constructed than spontaneous patient descriptions.

Tenth, the minimisation detection patterns have not been formally validated for sensitivity or specificity, and await participatory validation with native speakers of each of the 11 languages. Eleventh, the verification agent and uncertainty-aware routing mechanisms were implemented after the formal vignette validation and their impact on triage safety has not been independently assessed. Twelfth, WhatsApp-based delivery systematically excludes patients without smartphones, mobile data, or digital literacy – populations that may have the greatest need for healthcare navigation support. Finally, the system has not been tested in the context of South Africa’s electricity supply instability (load-shedding), although the DCSL’s independence from cloud AI provides architectural resilience. A single two-level under-triage event (V072, isiXhosa burns vignette) was observed in the reported validation; this is described in detail in Discussion. Burns classification without visual inspection is a known edge case for text-based triage, and addition of an ORANGE-level burns discriminator to the DCSL is a named pre-pilot priority.

## Conclusions

In a developer-led preliminary safety assessment spanning 120 SATS-aligned clinical vignettes across four languages, subsequently extended with a 121-vignette DCSL safety consistency evaluation across all 11 official South African languages and a 220-call framing sensitivity evaluation, BIZUSIZO achieved an under-triage rate of 3.3% (4/120) with no RED emergencies missed within the primary validation vignette set, and consistent detection of RED emergencies across all 11 official languages in structured regression testing scenarios. A single two-level under-triage event on a non-RED presentation (V072, isiXhosa burns vignette) was observed and is discussed in detail above as a specific architectural gap in ORANGE-level DCSL coverage. The DCSL correctly identified all life-threatening emergencies independent of AI within the structured test suite. In the framing sensitivity evaluation, the full pipeline achieved 95.0% RED invariance under adversarial framing compared to 50.9% for AI-only classification. Language-specific ceiling effects identified in Xitsonga and Tshivenda during post-validation 121-vignette regression testing of the Haiku 4.5 configuration were resolved by upgrading the production AI classifier to Claude Sonnet 4 prior to pilot deployment; formal validation of the Sonnet 4 production configuration on an independent held-out vignette set is planned as a prerequisite for pilot commencement.

The primary limitation of this work is its developer-led design: the same author who built the system designed the vignettes and assigned the gold standard. Every positive finding must be interpreted in that light. The necessary and sufficient next step is a prospective pilot concordance study in which BIZUSIZO’s pre-arrival triage is compared against independent nurse triage at arrival, using real patient presentations, with gold standard assignment by a panel of clinicians who had no role in the system’s development.

South Africa’s NHI reform creates an implementation pathway for pre-arrival triage infrastructure that did not previously exist. Whether BIZUSIZO or any comparable system can reduce unnecessary emergency attendance, improve chronic medication routing, and generate the longitudinal patient navigation data the NHI requires – without concentrating clinical risk in a single AI point of failure – is an empirical question that only prospective deployment can answer. The safety architecture described here – specifically, deterministic override of life-threatening presentations combined with automated regression testing and framing sensitivity evaluation as deployment gates – may represent a generalisable approach to safe deployment of AI-assisted triage in resource-constrained settings. Whether this architecture translates into improved clinical outcomes and system efficiency remains to be established through prospective evaluation. More broadly, this work proposes that AI triage safety in resource-constrained settings is most productively framed as a systems engineering problem – solved through layered deterministic constraints, multi-agent verification, and continuous monitoring – rather than a model accuracy problem.

## Supporting information

Additional file 2: DECIDE-AI reporting checklist

Additional file 3: DCSL discriminator catalogue

Additional file 4: March 2026 validation correction note

## Data Availability

The complete set of 120 clinical vignettes with gold standard triage assignments and the validation results are available from the corresponding author on reasonable request. The clinical rules engine source code is available at github.com/Bongekilenkosi/bizusizo.

## List of abbreviations

AI: artificial intelligence
API: application programming interface
ART: antiretroviral therapy
BSA: body surface area
CCMDD: Central Chronic Medicine Dispensing and Distribution
CI: confidence interval
CI/CD: continuous integration / continuous deployment
DCSL: Deterministic Clinical Safety Layer
DoH: Department of Health
EMS: emergency medical services
FHIR: Fast Healthcare Interoperability Resources
GPS: Global Positioning System
HREC: Human Research Ethics Committee
IQR: interquartile range
LLM: large language model
LOC: loss of consciousness
NHI: National Health Insurance
NICD: National Institute for Communicable Diseases
PHC: primary healthcare
PII: personally identifiable information
POPIA: Protection of Personal Information Act
REF: referral identifier
SATS: South African Triage Scale
SOP: standard operating procedure
TB: tuberculosis
TEWS: Triage Early Warning Score

## Declarations

### Ethics approval and consent to participate

This study did not involve human participants and did not collect, process, or analyse any identifiable patient data. The vignette-based evaluation used fictional clinical scenarios adapted from published SATS discriminators; no clinical intervention was administered. Under the South African Department of Health Ethics in Health Research: Principles, Processes and Structures (2nd edition, 2015), research not involving human participants or identifiable personal information falls outside the scope of mandatory Research Ethics Committee review. On this basis, formal review by the University of the Witwatersrand Human Research Ethics Committee (Medical) was not sought for this vignette-only technical safety evaluation, and no ethics committee reference number applies to this study. Confirmation of exemption will be obtained from the Wits HREC (Medical) should the journal or reviewers request it. The Clinical Governance Lead (SP, registered nurse) provided independent blinded verification of the gold-standard triage assignments in her professional capacity as a named team member of the system under evaluation; her ratings were provided as part of system development and governance rather than as a research subject. Formal ethics approval for the planned prospective patient pilot at three Tshwane primary healthcare facilities (Eersterus Community Health Centre, Skinner Clinic, and Soshanguve Community Health Centre) will be obtained from the Wits HREC (Medical) before any patient data are collected.

### Consent for publication

Not applicable.

### Availability of data and materials

The 120 primary validation vignettes with gold standard triage assignments and the DCSL source code are available from the corresponding author on request. The system architecture, validation pipeline, and 121-vignette DCSL multilingual regression test suite are publicly available at the project code repository (github.com/Bongekilenkosi/BIZUSIZO). Regression test results from each deployment are stored in a structured database with full timestamps. Primary validation scoring data will be made available upon reasonable request following pilot completion. The following additional files accompany this manuscript: Additional file 1 (Supplementary Table S1; .xlsx) contains the complete 120-vignette dataset sourced from the 1 April 2026 Haiku re-run (vignette_results_Apr2026.json), including patient-register symptom text in original language with English translations, reference-standard triage assignments, independent blinded nurse ratings, and per-vignette AI and final triage classifications. Additional file 2 (DECIDE-AI checklist; .pdf) contains the completed reporting checklist for the early-stage clinical evaluation of the system. Additional file 3 (DCSL discriminator catalogue; .pdf) contains the complete list of 53 clinical discriminator categories with representative keyword patterns across all 11 official languages. Additional file 4 (March 2026 validation correction note; .pdf) documents the reconciliation between the initial 30–31 March 2026 validation and the 1 April 2026 preserved re-run that is the primary validation reported in this manuscript.

### Authors’ contributions

BN conceived and designed the study, developed the BIZUSIZO system, designed the clinical vignettes, conducted the validation testing, analysed the results, and wrote the manuscript. The author read and approved the final manuscript.

## Acknowledgements

The author thanks Sheila Plaatjie, RN, for clinical governance review; Prof Tobias Chirwa, PhD, School of Public Health, University of the Witwatersrand, for agreeing to serve as Co-Principal Investigator of the planned EVAH pilot study; Anele Khuzwayo, MBA, for financial and operational guidance; and Ayanda Nkosi, LLB candidate, for legal and regulatory review.

