## Additional file 2: DECIDE-AI reporting checklist for "Design and preliminary safety validation of a hybrid deterministic–AI triage system for multilingual primary healthcare: a WhatsApp-based vignette study in South Africa"

**Reporting standard:** DECIDE-AI (Developmental and Exploratory Clinical Investigations of Decision-support systems driven by Artificial Intelligence), Vasey et al., *BMJ* 2022;377:e070904.

**Note on applicability:** DECIDE-AI is designed for early-stage *live clinical evaluation* of AI-based decision support systems. The present study is a **pre-live vignette-based technical safety validation** conducted prior to the prospective patient-facing pilot. Items that presuppose live patient interaction (e.g., real patient outcomes, live user-interface evaluation) are marked *"To be reported in the prospective pilot study; design specified in this manuscript"* rather than omitted. This follows the DECIDE-AI authors' guidance that pre-live safety evaluations may report the subset of items relevant to the study stage while specifying the forward plan for remaining items.

---

---

### AI-specific items (1a–10)

#### Item 1a — Identification of the study as involving an AI decision-support system

**Reported:** Title (*"hybrid deterministic-AI triage system"*), abstract, Keywords (*AI-assisted triage*). Background §4.

#### Item 1b — Intended use and claimed advantage

**Reported:** Background §7 (three specific contributions); Methods: Clinical governance framework (*"positioned as a clinical decision support and navigation tool, not a medical device"*). Intended use: pre-arrival triage for self-presenting patients in South African primary healthcare, WhatsApp-delivered, multilingual, SATS-aligned.

#### Item 2 — Explanation of algorithmic methods (model type, training data, validation data, inputs, outputs, version)

**Reported:** Methods — "System architecture and patient journey" (Claude Haiku 4.5, model identifier `claude-haiku-4-5-20251001`, temperature 0.1, used as the free-text classifier for the 120-vignette validation reported; Claude Sonnet 4, `claude-sonnet-4-20250514`, in current production pending independent held-out validation). Methods — "Clinical methodology: SATS alignment" (system prompt, confidence-score output, 50-word reasoning trace, TEWS proxy). Methods — "Deterministic safety architecture" (case-insensitive substring matching; no fuzzy matching, NLP, or ML; ~960 keyword patterns across 53 discriminators × 11 languages). Training data: not applicable — no fine-tuning was performed; the commercial Anthropic API was used as-is. Input: free-text symptom description in any of 11 official South African languages. Output: RED/ORANGE/YELLOW/GREEN with confidence score 0–100.

#### Item 3 — Patient and public involvement in system design

**Reported:** Methods — "Multilingual architecture" (human translators for safety-critical messages), "Deterministic safety architecture" (native-speaker review pending for Sepedi, Setswana, Xitsonga, Tshivenda, isiNdebele). Limitations: formal PPI not conducted at design stage; acknowledged as a gap. Prospective pilot will include community advisory input via the Clinical Advisory Board.

#### Item 4 — Human factors considerations (user interface, training, integration into workflow)

**Reported:** Methods — "System architecture and patient journey" steps 1–8; Figure 1; Methods — "Clinical governance framework" (structured nurse feedback loop: Agree/Disagree, staff identifier, audit trail). Formal usability evaluation to be reported in the prospective pilot study; design specified in this manuscript.

#### Item 5 — Eligibility criteria of study participants

**Not applicable** — no human participants in the present vignette-based technical safety validation. Eligibility for the prospective pilot: self-presenting patients at three Tshwane primary healthcare facilities (Eersterus CHC, Skinner Clinic, Soshanguve CHC); full eligibility protocol to be reported in the pilot study manuscript and submitted to Wits HREC (Medical) prior to patient-facing deployment.

#### Item 6 — Study setting and its relevance to the intended use

**Reported:** Background §1–2 (South African public primary healthcare; approximately 3,900 clinics; 83% of population; long waiting times; CCMDD-dominant prescription load); Background §4 (absence of pre-arrival digital triage across 11 official languages). Methods — "System architecture and patient journey" (WhatsApp delivery rationale; 96.1% household mobile-phone penetration; 68% WhatsApp active-user share).

#### Item 7 — Version control, software updates, system maintenance during the study

**Reported:** Methods — "Deterministic safety architecture" (DCSL trigger patterns frozen prior to formal validation; validation vignettes not reused in CI regression testing; CI pipeline blocks deployment on any safety failure in the 121-vignette multilingual regression suite). Methods — "Post-hoc framing sensitivity evaluation" (surfaced coverage gaps permitted to feed into the CI pipeline but not retrospectively into validation). Model identifier freeze: `claude-haiku-4-5-20251001` for the reported validation; any change to the production classifier (e.g., the subsequent upgrade to Sonnet 4) flagged as requiring held-out revalidation.

#### Item 8 — Human oversight of the AI system

**Reported:** Methods — "Multi-agent verification for high-acuity classifications" (secondary AI verification, disagreement routed to nurse review on clinical dashboard, no automatic downgrade of high-acuity classifications). Methods — "Low-confidence uncertainty-aware routing" (confidence <75% → patient advisory message + nurse review flag, classification preserved). Methods — "Clinical governance framework" (monthly clinical audits of 40 randomly selected triage conversations; Clinical Governance Lead is a registered nurse).

#### Item 9 — Identification and handling of AI system errors, failures, and unintended consequences

**Reported:** Methods — "Deterministic safety architecture" (DCSL-as-override architecture is the primary error-handling mechanism; independent of AI availability); Methods — "Symptom completeness validation" (architectural response to Bean et al. 2024 real-world input-incompleteness failure mode); Methods — "Runtime minimisation detection" (architectural response to cultural minimisation patterns). Results — "Stress testing findings" (three critical DCSL bugs discovered pre-validation: `can't breathe` ≠ `shortness of breath`; isiXhosa–isiZulu breathing-verb mismatch; `bitten by a snake` ≠ `snake bite`; all corrected and added to CI). Results — "Post-hoc framing sensitivity evaluation" (quantifies and contains a specific AI failure mode via DCSL rescue; 18 of 23 AI drift cases rescued). Residual risks named in Limitations: vocabulary-completeness dependency, real-world misspellings, novel colloquialisms.

#### Item 10 — Data security, privacy, and regulatory compliance

**Reported:** Background §3 (NHI Act 20 of 2023 context); Abstract and Cover letter (POPIA compliance framework: informed consent available in 11 languages, data minimisation, PII stripping prior to external API calls, full audit logging). Methods — "System architecture and patient journey" step 2 (consent includes AI-use disclosure, POPIA reference, national emergency number). STOP command logged as formal consent withdrawal. DPIA v2.7 complete (`docs/dpia_v2.7_complete.md`). Data stays in Supabase; no patient-identifiable data sent to external APIs. Regulatory positioning: clinical decision support and navigation tool, not a SAHPRA-regulated medical device; SAHPRA classification assessment on file (`docs/sahpra_classification_assessment.md`).

---

---

### Generic reporting items (11–17)

#### Item 11 — Abstract (structured, includes key performance measures and uncertainty)

**Reported:** Abstract includes Background, Methods, Results (with point estimates and 95% confidence intervals for under-triage, over-triage, weighted kappa), and Conclusions. Keywords include safety-relevant terms.

#### Item 12 — Background (clinical need, current practice, AI rationale)

**Reported:** Background §§1–7. Clinical need: waiting times, CCMDD prescription load, pre-arrival routing gap. Current practice: SATS is hospital-validated; no 11-language pre-arrival digital triage. AI rationale: structured safety architecture with AI as one of three pathways, not the primary safety mechanism.

#### Item 13 — Study design (prospective/retrospective, comparison, blinding)

**Reported:** Methods — "Preliminary safety assessment: vignette-based testing" (developer-led technical safety validation; gold-standard reference rated by author with independent blinded verification by registered nurse as Clinical Governance Lead; discrepancies quantified with inter-rater kappa = 0.678, 95% CI 0.577–0.763). Study positioned explicitly as pre-live safety validation, not clinical effectiveness study. Post-hoc framing sensitivity evaluation reported separately and labelled as post-hoc.

#### Item 14 — Sample size rationale

**Reported:** Methods — "Preliminary safety assessment: vignette-based testing" (120 vignettes selected to provide 30 cases per language across four languages, with level coverage 20 RED / 32 ORANGE / 44 YELLOW / 24 GREEN; sizing consistent with published SATS vignette validation studies using 50 and 42 vignettes). Wide confidence intervals on primary outcomes explicitly discussed in Results.

#### Item 15 — Outcomes (primary, secondary, how measured)

**Reported:** Methods — "Preliminary safety assessment" (primary: under-triage rate; secondary: over-triage rate, exact concordance, agreement within one SATS level, quadratic weighted Cohen's kappa, composite safety score). Methods — "Post-hoc framing sensitivity evaluation" (primary: RED invariance under adversarial framing; DCSL rescue count as secondary).

#### Item 16 — Analysis (statistical methods, handling of missing data)

**Reported:** Methods — "Preliminary safety assessment" (Clopper-Pearson exact binomial confidence intervals selected for small sample and low event rate; quadratic weighted Cohen's kappa as primary agreement metric; composite safety score flagged as study-specific ordinal weighting, not a standard measure). Missing data: one blinded-nurse rating was missing; kappa calculated on

119 evaluable vignettes and this is stated explicitly.

#### Item 17 — Ethics and consent

**Reported:** Cover letter and Methods — "Preliminary safety assessment" (no human participants; no identifiable patient data; vignettes drafted against published SATS discriminators; 2015 DoH Ethics in Health Research guidelines place non-human-participant research outside mandatory REC review; Wits HREC (Medical) review will be obtained before the prospective patient pilot commences).

---

---

### Additional items recommended by DECIDE-AI for AI studies

#### Subgroup performance

**Reported:** Methods — "Clinical governance framework" (subgroup performance reporting disaggregated by language, facility, age group, and sex; automated disparity flags in the governance dashboard). Results — Performance by triage level (per-level disaggregation reported). Per-language kappa to be reported in the prospective pilot (the 120-vignette sample size supports level disaggregation but is underpowered for fully disaggregated per-language kappa).

#### Calibration and confidence

**Reported:** Results — "Overall system performance" (98% of AI classifications exceeded the 75% confidence threshold; relationship between AI confidence and classification behaviour discussed — *"high AI confidence did not preclude over-triage"*). Methods — "Low-confidence uncertainty-aware routing" specifies behaviour for the 2% low-confidence cases.

#### Interpretability / explainability

**Reported:** Methods — "Clinical methodology: SATS alignment" (AI returns a structured reasoning trace of up to 50 words identifying the SATS step that determined the classification; logged for audit). Methods — "Deterministic safety architecture" (DCSL is fully interpretable by design: every RED override returns a named rule identifier with 100% confidence).

#### Model drift and ongoing monitoring

**Reported:** Methods — "Deterministic safety architecture" (121-vignette CI regression suite as deployment gate across all 11 languages). Methods — "Clinical governance framework" (monthly clinical audits). Future prospective pilot will add 24-hour and 72-hour patient-reported outcome linkage.

#### Economic and resource implications

**Not reported in this manuscript; to be addressed in the planned pilot concordance and implementation study.**

---

**Checklist completed:** 20 April 2026, Bongekile Esther Nkosi-Mjadu
