## Additional file 3: DCSL discriminator catalogue for "Design and preliminary safety validation of a hybrid deterministic–AI triage system for multilingual primary healthcare: a WhatsApp-based vignette study in South Africa"

### Additional file 3: DCSL discriminator catalogue — 11-language coverage matrix

**Source of truth:** `lib/triage.js`, function `applyClinicalRules`. This catalogue is generated programmatically from the production source code via `scripts/extract_dcsl_catalogue.js`, ensuring the manuscript's claimed discriminator set matches what is actually deployed. Language coverage is determined by semantic scanning of each rule's keyword list against per-language marker dictionaries (characteristic phonemes and vocabulary for each of the 11 official South African languages), not by rule-ID suffix.

**Languages:**

| Code | Language |
|---|---|
| **en** | English |
| **zu** | isiZulu |
| **xh** | isiXhosa |
| **af** | Afrikaans |
| **nso** | Sepedi |
| **tn** | Setswana |
| **st** | Sesotho |
| **ts** | Xitsonga |
| **ss** | siSwati |
| **ve** | Tshivenda |
| **nr** | isiNdebele |

**Reading the table:** each row is one DCSL rule identifier. `Y` indicates the rule's keyword list contains at least one marker characteristic of that language (i.e., the rule can fire on a patient message written in that language); `·` indicates no detected coverage. English (en) is always `Y` because rule triggers are authored in English and language-specific keywords are additions on top.

**Validation status:** Keyword sets for en, zu, xh, af are the formally validated vocabulary used in the 120-vignette validation reported in the manuscript. Keyword sets for nso, tn, st, ts, ss, ve, nr were added to the codebase prior to pilot and await independent review by native speakers (see `docs/DCSL_Native_Speaker_Review.md`). The 121-vignette multilingual DCSL regression suite passes on all 11 languages for the RED discriminators it tests.

##### RED discriminators (16 distinct base rule identifiers)

| Base rule ID | en | zu | xh | af | nso | tn | st | ts | ss | ve | nr |
|---|---|---|---|---|---|---|---|---|---|---|---|
| acs_radiation | Y | Y | · | Y | Y | Y | Y | Y | Y | Y | · |
| active_seizure | Y | Y | Y | Y | Y | Y | Y | Y | Y | Y | Y |
| anaphylaxis | Y | Y | · | Y | Y | Y | Y | Y | Y | Y | · |
| cardiac_emergency | Y | Y | Y | Y | Y | Y | Y | Y | Y | Y | Y |
| cardiac_emergency_radiation | Y | Y | · | Y | Y | Y | Y | Y | Y | Y | Y |
| envenomation | Y | Y | Y | Y | Y | · | Y | Y | Y | Y | · |
| meningococcal_rash | Y | Y | · | · | · | · | · | · | · | · | · |
| neonatal_apnoea | Y | Y | Y | Y | Y | · | Y | · | Y | Y | Y |
| obstetric_cord_or_fetal | Y | Y | Y | Y | · | Y | · | · | Y | Y | · |
| obstetric_haemorrhage | Y | Y | · | Y | Y | · | Y | Y | Y | Y | Y |
| paediatric_unconscious | Y | Y | Y | Y | Y | Y | Y | · | Y | Y | Y |
| respiratory_cardiac_arrest | Y | Y | Y | Y | Y | Y | Y | Y | Y | Y | Y |
| severe_burns | Y | Y | Y | Y | Y | Y | Y | Y | Y | Y | Y |
| severe_burns_context | Y | Y | Y | Y | Y | Y | Y | Y | Y | Y | Y |
| traumatic_haemorrhage | Y | Y | · | Y | · | · | · | · | · | Y | · |
| unconscious | Y | Y | Y | Y | Y | Y | Y | Y | Y | Y | Y |

##### ORANGE discriminators (22 distinct base rule identifiers)

| Base rule ID | en | zu | xh | af | nso | tn | st | ts | ss | ve | nr |
|---|---|---|---|---|---|---|---|---|---|---|---|
| acute_abdomen | Y | Y | Y | Y | Y | Y | Y | Y | Y | Y | Y |
| acute_confusion_chronic | Y | Y | · | Y | Y | Y | Y | Y | Y | Y | Y |
| acute_confusion_dm | Y | · | · | Y | · | · | · | · | · | · | · |
| burns | Y | Y | Y | · | · | · | · | · | · | · | · |

| | | | | | | | | | | | |
|---|---|---|---|---|---|---|---|---|---|---|---|
| burns_significant | Y | Y | Y | Y | Y | Y | Y | Y | Y | Y | Y |
| ectopic_pregnancy | Y | Y | Y | Y | · | · | · | · | · | · | Y |
| febrile_seizure | Y | Y | · | Y | Y | · | Y | Y | Y | Y | · |
| head_trauma_loc | Y | Y | Y | Y | Y | Y | Y | Y | Y | Y | Y |
| high_energy_mechanism | Y | Y | Y | Y | Y | Y | Y | Y | Y | Y | Y |
| hiv_meningism | Y | Y | · | Y | Y | Y | Y | Y | Y | Y | · |
| infant_sepsis_screen | Y | Y | · | Y | Y | · | Y | Y | Y | Y | · |
| open_fracture | Y | Y | Y | Y | Y | Y | Y | Y | Y | Y | Y |
| post_ictal | Y | Y | · | · | Y | Y | Y | Y | Y | Y | Y |
| pre_eclampsia | Y | Y | Y | Y | Y | Y | Y | Y | Y | Y | · |
| preterm_labour | Y | Y | · | Y | · | · | · | · | Y | · | · |
| psychiatric_emergency_imminent | Y | Y | · | Y | Y | Y | Y | Y | Y | Y | Y |
| severe_asthma | Y | Y | · | · | · | · | Y | Y | Y | · | · |
| severe_hypoglycaemia | Y | Y | · | Y | Y | Y | Y | Y | Y | Y | Y |
| stroke_arm_weakness | Y | Y | · | Y | Y | Y | Y | Y | · | Y | · |
| stroke_facial_droop | Y | Y | Y | Y | Y | Y | Y | · | · | · | · |
| stroke_speech | Y | Y | Y | Y | Y | Y | Y | Y | · | Y | Y |
| thunderclap_headache | Y | Y | Y | Y | Y | Y | Y | Y | Y | Y | · |

##### YELLOW discriminators (20 distinct base rule identifiers)

| Base rule ID | en | zu | xh | af | nso | tn | st | ts | ss | ve | nr |
|---|---|---|---|---|---|---|---|---|---|---|---|
| abuse_assault | Y | Y | Y | Y | · | · | · | · | Y | · | · |
| appendicitis_pattern | Y | · | · | · | · | · | · | · | · | · | · |
| asthma_inhaler_failure | Y | · | · | · | · | · | · | · | · | · | · |
| deep_wound | Y | · | · | · | · | · | · | · | · | · | · |
| dka | Y | Y | · | Y | Y | Y | Y | Y | Y | Y | Y |
| eye_emergency | Y | · | · | · | · | · | · | · | · | · | · |
| gi_bleeding | Y | Y | · | Y | · | · | · | · | · | Y | · |
| hiv_fever | Y | · | · | · | · | · | · | · | · | · | · |
| hypertensive_urgency | Y | Y | Y | Y | Y | Y | Y | Y | Y | Y | Y |
| hypertensive_urgency_reading | Y | · | · | · | · | · | · | · | · | · | · |
| lower_abdo_missed_period | Y | · | · | · | · | · | · | · | · | · | · |
| meningism | Y | Y | · | Y | Y | · | Y | Y | · | Y | · |
| possible_fracture | Y | · | · | · | · | · | · | · | · | · | · |
| pregnancy_complication | Y | Y | · | Y | · | · | · | · | Y | · | · |
| pyelonephritis | Y | Y | · | Y | · | · | · | · | · | · | · |
| severe_dehydration_vulnerable | Y | · | · | · | · | · | · | · | · | · | · |
| severe_pain | Y | Y | Y | Y | · | · | · | · | · | · | · |
| suicidal_ideation | Y | Y | · | · | · | · | · | · | · | · | · |
| tb_triad | Y | Y | · | · | · | · | · | · | · | · | · |
| testicular_torsion | Y | · | · | · | · | · | · | · | · | · | · |

---

#### Summary

**Live code (rule identifier count, extracted 2026-04-20):**

- **RED:** 16 distinct base rule identifiers
- **ORANGE:** 22
- **YELLOW:** 20
- **Total:** 58

**Per-language coverage (number of rule identifiers with keywords for each language):**

| Level | en | zu | xh | af | nso | tn | st | ts | ss | ve | nr |
|---|---|---|---|---|---|---|---|---|---|---|---|
| RED | 16/16 | 16/16 | 10/16 | 15/16 | 13/16 | 11/16 | 13/16 | 11/16 | 14/16 | 15/16 | 10/16 |
| ORANGE | 22/22 | 21/22 | 11/22 | 19/22 | 17/22 | 15/22 | 18/22 | 17/22 | 16/22 | 16/22 | 11/22 |
| YELLOW | 20/20 | 10/20 | 3/20 | 8/20 | 3/20 | 2/20 | 3/20 | 3/20 | 4/20 | 4/20 | 2/20 |

**Manuscript claim reconciliation.** The manuscript states *“53 clinical discriminator categories (14 RED, 19 ORANGE, 20 YELLOW)”*. The code implements 58 rule identifiers (16 RED / 22 ORANGE / 20 YELLOW), of which 5 are variant patterns of base categories and 2 (`infant_sepsis_screen`, `hiv_meningism`) are post-validation safety-net rules added after the 120-vignette validation. The 53-category count in the manuscript refers to the base taxonomy at the time of validation; see manuscript footnote on Table 1 for the reconciliation.

*Generated:* 2026-04-20 via `node scripts/extract_dcsl_catalogue.js`
