## Additional file 4: March 2026 validation correction note for "Design and preliminary safety validation of a hybrid deterministic–AI triage system for multilingual primary healthcare: a WhatsApp-based vignette study in South Africa"

### Additional file 4: March 2026 validation — data provenance correction record

**Status:** Retrospective correction, written 20 April 2026 during medRxiv resubmission preparation. This is the canonical in-repo record of the divergence between the manuscript's originally reported headline numbers (95 correct / 22 over-triaged / 3 under-triaged / 0 two-level over-triage) and the preserved validation data on disk.

The manuscript is being updated to report the preserved April re-run numbers. This note explains why, and documents the provenance of each dataset.

---

#### 1. Summary of the correction

- **Originally reported (in manuscript, BMC cover letter, `BIZUSIZO_Vignette_Validation_Results .pdf` dated 31 March 2026):** 2.5% under-triage (3/120); 79.2% concordance (95/120); 18.3% over-triage (22/120); weighted κ=0.889; 0 two-level over-triage.
- **Corrected values (from `vignette_results_Apr2026.json`, timestamp 2026-04-01T22:42):** 3.3% under-triage (4/120); 80.0% concordance (96/120); 16.7% over-triage (20/120); weighted κ=0.891 (bootstrap 95% CI 0.827–0.932); 1 two-level over-triage; 1 two-level under-triage.
- **Under-triage cases, corrected:** V041, V042, V072, V103 (previously reported as V041, V072, V103 — V042 was omitted from the count).

---

#### 2. What happened

##### 2.1 The 30 March 2026 Haiku validation run

The 120-vignette validation was run on Claude Haiku 4.5 (`claude-haiku-4-5-20251001`) at 23:52 on 30 March 2026. The raw output is preserved in two locations:

- `C:\Users\bonge\OneDrive\Desktop\Stress Testing\vignette_results_Mar2026` (36,918 bytes, file mtime 2 April 2026 01:35)
- `C:\Users\bonge\Downloads\vignette_results_Mar2026.json` (36,187 bytes)
- `vignette_export.json` in the present repository (identical internal content; timestamp `2026-03-30T23:52:21.932Z`)

This run produced five parse errors on vignettes **V050, V065, V070, V072, V073** (isiXhosa tokenisation issue, resolved in subsequent tool hardening). On the 115 successfully scored cases, outcomes were:

- 93 correct / 19 over-triaged / 3 under-triaged (V041, V042, V103)

##### 2.2 The 31 March 2026 validation document

On the night of 30–31 March 2026, after the 23:52 run produced parse errors on V050, V065, V070, V072, V073, I retried only those five cases. I did not re-run any other vignette. I did not revise any gold-standard assignment. The retry outputs were used to produce the headline totals in the 31 March 01:00 validation PDF (`BIZUSIZO_Vignette_Validation_Results .pdf`), but the retry output was not saved to disk as a dated JSON file.

The PDF's headline totals (95 correct / 22 over-triaged / 3 under-triaged / 0 two-level over-triage) combine the 115 scored cases from the preserved 30 March raw run (93 correct / 19 over / 3 under) with the 5 retry outputs. Reconciling these totals arithmetically requires the retry outputs to have been 2 CORRECT and 3 OVER-TRIAGED — consistent with the DCSL rule behaviour expected on each vignette text but not independently verifiable against a preserved retry file.

The PDF's under-triage narrative names V041, V072, V103. This is inconsistent with the preserved 30 March raw run, which shows V041, V042, V103 as the three under-triage cases among the 115 successfully scored. V042 was under-triaged and omitted from the PDF narrative; V072 was a parse error in the preserved raw run and was scored as under-triaged only after the retry. The retry arithmetic produces 2 correct + 3 over, which would mean no new under-triage cases from the retry — but V072's retry output is not preserved, so whether it was scored correct/over/under in the retry cannot be independently verified from disk.

**Because the retry output was not preserved, the PDF's 95/22/3/0 totals are not independently reproducible from any preserved data file.** This is the load-bearing reason the manuscript is being corrected to the 1 April re-run numbers (`vignette_results_Apr2026.json`), in which all 120 cases scored successfully with no parse errors and the data is preserved in full.

##### 2.3 The 1 April 2026 Haiku re-run

On 1 April 2026, medRxiv returned the initial submission requesting the vignette dataset as supplementary data for preprint posting. In response, a full 120-vignette re-run of Claude Haiku 4.5 was conducted at 22:42 on 1 April 2026 and preserved as:

- `C:\Users\bonge\OneDrive\Desktop\Stress Testing\vignette_results_Apr2026.json` (35,730 bytes, internal timestamp `2026-04-01T22:42:11.581Z`)

This re-run produced 120/120 successfully scored results (zero parse errors) and is the preserved validation dataset. Outcomes:

- 96 correct / 20 over-triaged / 4 under-triaged / 1 two-level over-triage / 1 two-level under-triage
- Under-triage cases: V041 (isiZulu, ORANGE→YELLOW), V042 (isiZulu, ORANGE→YELLOW), V072 (isiXhosa, ORANGE→GREEN), V103 (Afrikaans, ORANGE→YELLOW)
- Two-level under-triage: V072 (ORANGE→GREEN, 2-level) — a burns presentation that the classifier judged as localised thermal injury manageable at PHC level
- Two-level over-triage: V054 (English deep laceration, YELLOW→RED) — surfaced as a post-validation rule-engine-expansion artefact when keyword coverage was broadened; this is subsequently documented as CI-regression-pipeline output

The 1 April re-run is fully reproducible from the preserved JSON file and an analysis script (`scripts/compute_apr1_metrics.js` in this repository).

##### 2.4 Inter-run variance between 30 March and 1 April Haiku runs

Comparing the 30 March raw run (on the 115 successfully scored cases) to the 1 April re-run (same cases, same model, same prompt, same `temperature=0.1`), classification differed on approximately 6/120 cases (~5%). This stochastic variation is expected behaviour for a large language model at non-zero temperature, and is reported as a reproducibility observation — not as evidence of tool modification. The DCSL trigger patterns and SATS-alignment system prompt were unchanged between the two runs.

##### 2.5 The 2 April 2026 Sonnet 4 production upgrade

Following Haiku validation, the BIZUSIZO production pipeline was upgraded to Claude Sonnet 4 (`claude-sonnet-4-20250514`) on 2 April 2026 to resolve language-specific ceiling effects observed in Xitsonga and Tshivenda during post-validation regression testing. The Sonnet 4 full-run output is preserved at:

- `C:\Users\bonge\OneDrive\Desktop\Stress Testing\vignette_results_Sonnet_Apr2026.json` (internal timestamp `2026-04-02T00:31:50Z`)

The Sonnet 4 configuration is disclosed in the manuscript as a **post-validation production change**, not as an independently validated classifier. Sonnet 4 numbers are explicitly *not* substituted into the validation results section; the manuscript reports Haiku 4.5 validation and discloses the Sonnet upgrade as a limitation pending formal held-out revalidation. This framing avoids hypothesising-after-results-are-known (HARKing) and was established earlier in this preparation cycle as the correct methodological position.

---

#### 3. Why this correction is being made now, not earlier

The discrepancy between the 31 March PDF headline numbers and the 1 April preserved re-run was identified on 20 April 2026 during medRxiv resubmission preparation, when the supplementary table was built from `vignette_results_Apr2026.json` and the per-case outcomes did not match the manuscript's named under-triage cases (V041, V072, V103). Verification of the underlying files disclosed that:

- The 30 March raw run was preserved and shows V042 as an under-triage case (gold ORANGE, system YELLOW) omitted from the PDF's narrative
- The 1 April re-run confirms V042 as an under-triage case and adds V072 as a 2-level under-triage (gold ORANGE, system GREEN)
- No intermediate scoring file exists that would reconcile the PDF's 95/22/3/0 totals to any preserved data

Making the correction now, before medRxiv resubmission, is the honest path. Reporting numbers that cannot be reproduced from preserved data, while preserved data on disk shows different values, would be a reproducibility failure regardless of whether the earlier numbers were defensible at the time they were written.

---

#### 4. Impact on the manuscript's conclusions

The corrected numbers do not change the paper's core safety claim:

- Under-triage remains well below the published South African SATS benchmark range (9% Zithulele in-hospital [14], 29.5% prehospital EMS [10]).
- No RED under-triage was observed; all four under-triage cases are ORANGE misclassified to YELLOW or GREEN.
- Over-triage is now lower (16.7% vs 18.3%) — still within the published SA SATS range.
- Concordance is slightly higher (80.0% vs 79.2%).
- Weighted κ is slightly higher (0.891 vs 0.889).
- Rules engine RED sensitivity: 100% (20/20), unchanged.
- The V054 two-level over-triage event becomes a documented instance of the continuous-integration pipeline surfacing a rule-expansion artefact — consistent with the paper's CI-as-safety-infrastructure contribution.
- The V072 two-level under-triage event (burns classified as GREEN) is reported as a safety-relevant observation requiring explicit Discussion coverage: the AI's reasoning was clinically coherent (localised thermal injury manageable at PHC level) but reflects a known limitation of text-only triage without visual burn assessment, appropriate for acknowledgement in Limitations.

---

#### 5. Actions taken

- **Reverted:** the three draft corrigenda committed as `b04fe3e` on 20 April 2026 (retrospective adjudication note, PDF corrigendum, Methods paragraph draft) — these were built on a partial understanding before the preserved 1 April re-run was located.
- **This document:** the canonical in-repo record of the correction.
- **`docs/validation_document_v072_correction.md`:** to be rewritten in a separate commit to remove the previously incorrect attribution to the 8 April re-run and replace with the correct attribution to the 1 April re-run.
- **Supplementary Table S1:** to be regenerated from `vignette_results_Apr2026.json` so every row is traceable to preserved data.
- **Manuscript and cover letter:** to be updated in a focused editing pass once this correction record is signed off.

---

#### 6. Author sign-off

**This record is accurate to the author's memory as of 20 April 2026 and describes what happened with the vignette validation data.** Sections 2.1, 2.3, 2.4, and 2.5 are directly verifiable from preserved JSON files. Section 2.2 describes the 30–31 March retry of the five parse-error cases from direct memory; the retry output was not saved to disk, and the absence of preservation is itself the reason the manuscript is being corrected to the 1 April re-run numbers.

Signed: Bongekile Esther Nkosi-Mjadu, MPH Date: 20 April 2026

---

*This correction record is intended to be cited in the manuscript's Methods section as the canonical data-provenance record, and to be retained in the public repository accompanying the paper.*

---

### Appendix A: Correction notice — BIZUSIZO Validation Document v2.0 (March 2026)

**Document affected:** *BIZUSIZO AI Triage Validation Results — 120-Vignette Preliminary Safety Assessment | Four South African Languages | March 2026 | Version 2.0* (file `BIZUSIZO_Vignette_Validation_Results .pdf`, mtime 31 March 2026 01:00).

**Status:** Correction identified 2026-04-20 during medRxiv resubmission preparation. Superseded by the canonical record above, which is the in-repo record of the full data-provenance correction. This appendix is retained as a scoped corrigendum to the external PDF and cross-references the canonical note.

**Previous version of this file:** before 2026-04-20, this document incorrectly attributed the PDF's headline numbers (2.5% / 79.2% / κ=0.889) to an 8 April Haiku post-DCSL-fix re-run. That attribution was wrong. The correct provenance is documented below and in the canonical correction note above.

---

---

#### A1. What was wrong in the Validation Document v2.0 PDF

Section 4 (Safety Interpretation), under the *"Under-triage (2.5%, 3/120)"* heading, the PDF stated:

> *The three under-triage cases were: V041 (isiZulu — head trauma with LOC, classified YELLOW), V072 (isiXhosa — significant burns, **classified GREEN**), and V103 (Afrikaans — acute confusion with diabetes, classified YELLOW).*

Two errors in this narrative:

1. **V042 omitted from the under-triage list.** The preserved 30 March 2026 raw Haiku run (`vignette_results_Mar2026` / `vignette_export.json`) shows V041, V042, V103 as the three under-triage cases among the 115 successfully scored vignettes. V042 (isiZulu — hot water scald to hands, ORANGE classified YELLOW) was an under-triage case and is absent from the PDF's narrative.
2. **V072 identified and labelled incorrectly.** V072 was a parse error in the preserved 30 March raw run, not a classified case. The PDF's identification of V072 as an under-triage case "classified GREEN" cannot be reconciled to the preserved 30 March data. It is a write-up error in the PDF narrative, not a reflection of what the preserved data shows.

---

#### A2. Provenance of the Validation Document v2.0 headline numbers

The PDF's headline totals (2.5% under-triage / 79.2% concordance / κ=0.889) were produced on 30–31 March 2026 by the author (BN) retrying the five parse-error cases (V050, V065, V070, V072, V073) after the 30 March 23:52 Haiku run, and combining those retry outputs with the 115 successfully scored cases to produce 95/22/3/0 totals. **The retry output was not saved to disk as a dated JSON file.** The PDF's totals therefore cannot be independently reproduced from any preserved data file.

The full data-provenance record is in the main body of this document above. That record describes the retry, the arithmetic reconciliation, and the reason the manuscript now reports 1 April 2026 re-run numbers (`vignette_results_Apr2026.json`) rather than the 31 March PDF's figures.

#### A3. What this means for the external Validation Document v2.0 PDF

The 31 March 2026 PDF should be treated as **a point-in-time development artefact** that predates the preserved 1 April Haiku re-run. It is not the source of the numbers now reported in the manuscript.

- **Do not re-issue the PDF as Version 2.1** with a minor transcription fix. The underlying data-provenance problem (unpreserved retry output) is more fundamental than a per-case label typo, and the cleanest path is to retire the PDF rather than incrementally patch it.
- **Superseded by:** `vignette_results_Apr2026.json` (1 April 2026 Haiku re-run, 120/120 scored, zero parse errors) as the preserved validation dataset, and the main body of this document as the provenance record.
- **If the PDF remains in circulation externally** (e.g., as an attachment to the 1 April 2026 EVAH submission), readers should be directed to the superseding records. An errata slip referencing this document can accompany any further circulation of the v2.0 PDF.

#### A4. What this correction does NOT do

- Does not alter anything in the preserved 1 April Haiku re-run (`vignette_results_Apr2026.json`) or the manuscript's corrected headline numbers (3.3% under-triage, 80.0% concordance, $\kappa$=0.891) drawn from that file.
- Does not introduce Sonnet 4 numbers into the validation reporting. Sonnet 4 remains disclosed in the manuscript as the post-validation production classifier, not as an independently validated alternative to Haiku 4.5.
- Does not re-open scope on the vignette set, the gold-standard assignments, or the SP blinded ratings. Those remain as they were at validation time.

#### A5. Cross-references

- Canonical correction record: the main body of this document (Additional file 4 of the manuscript submission package).
- Preserved 30 March raw run: `vignette_export.json` (in-repo) / `vignette_results_Mar2026` (in `Stress Testing/`) — 115/120 scored, 5 parse errors.
- Preserved 1 April re-run (primary validation dataset for the manuscript): `vignette_results_Apr2026.json`.
- Preserved 2 April Sonnet 4 post-validation configuration run: `vignette_results_Sonnet_Apr2026.json`.
- Supplementary Table S1 (rebuilt from 1 April Haiku re-run): `docs/supplementary_s1_vignettes.{csv,md}`.
